# Peer-guided movie neurofeedback to facilitate recovery in opioid use disorder

**DOI:** 10.64898/2026.09.18.26363414

**Authors:** Greg Kronberg, Natalie E. McClain, Maggie Boros, Shraeya Santhapuram, K. Rachel Drury, Augustin C. Hennings, Uri Hasson, Nelly Alia-Klein, Kenneth A. Norman, Rita Z. Goldstein

## Abstract

Real-time fMRI neurofeedback can modify clinically relevant brain states, but transferring brain dynamics between peers has not been tested, particularly in addiction, where social learning is central to recovery. Here we introduce peer-guided movie neurofeedback (PG-MovieNF), which trains participants to align their brain activity with treatment-related dynamics derived from peers further along in recovery viewing the same movie. Using longitudinal fMRI data from individuals with opioid use disorder watching Trainspotting before and after 8 weeks of adjunctive treatment, we derived a multivariate treatment-predictive signal in dorsolateral and dorsomedial prefrontal cortex. An independent cohort then received real PG-MovieNF or yoked sham feedback during the same film. Real feedback increased alignment with the peer-derived target signal, reduced craving and improved affective ratings. Target alignment was coupled to shared orbitofrontal cortex dynamics during real feedback and predicted craving reduction. Together, these findings establish proof of concept for PG-MovieNF to enhance addiction recovery.

## Introduction

Learning from others is essential to adaptive behavior. Across species, observation and imitation, and in humans, instruction and social feedback, allow individuals to acquire strategies that would be slow, costly or impossible to discover alone^1–3^. Psychology and psychiatry have long harnessed this principle through peer support, mutual-help groups and other lived-experience-based interventions^4–6^, improving patients’ well-being and treatment outcomes^7^. Successful social learning is associated with the construction of shared, context-dependent neural representations across people^8,9^, suggesting that these shared representations also support peer-based psychiatric interventions.

Substance use disorders provide a particularly relevant example of both the necessity and challenges of social learning in treatment. In drug addiction, drug-related cues and contexts acquire excessive salience, automatically capture attention, evoke craving, recruit habits, and weaken executive control^10,11^. These abnormalities span interacting prefrontal, striatal, insular and limbic systems rather than a single neural locus^10–13^. Recovery therefore likely requires more than improving any one function; it requires learning to process drug-related environments differently, reweighting salience and value, and engaging control in complex, cue-rich settings that typically evoke urges to use. Peer-guided recovery models (e.g., 12-step programs, group therapy) provide examples of how to navigate such drug-related environments from those with lived experience, harnessing the inherently social aspect of addiction that spans both active use and recovery^14–17^. However, translating another person’s recovery experience into one’s own internal state depends on neurocognitive functions often compromised in addiction, such as metacognition, mentalizing, insight and self-monitoring^12,18^. A brain-based intervention that conveys a peer’s recovery-related neural state directly, rather than through verbal communication, could therefore scaffold the process of social learning precisely as these impairments hinder it.

Real-time fMRI neurofeedback (rt-fMRI-NF) offers a more direct route for training recovery-relevant brain states by making ongoing brain activity available for learning^19,20^. In addiction, rt-fMRI-NF has shown promise for modulating cue-reactive brain responses and craving-related processes, but most studies have relied on single-region univariate signals, usually derived from the participant’s own brain and elicited by static cues^21^. More recently, rt-fMRI-NF has advanced beyond simple regional up- and down-regulation to the shaping of multivariate sensory patterns and distributed representational states^22–24^. Yet these approaches remain largely intra-individual, whereby the target state is derived from an individual’s own brain. The few efforts to derive target signals from an independent group^25–27^ have primarily trained static, image-derived categorical representations, but have not encompassed the rich temporal dynamics evoked by naturalistic stimuli such as movies or narratives. In part motivating the current study, there has been one rt-fMRI-NF study to date that delivered feedback in response to a naturalistic stimulus, whereby neural activity decoded from an independent group was used to bias a new group of listeners towards one of two interpretations of an ambiguous spoken narrative, in the general population^28^. This prior work establishes that distributed brain states can be shaped in real time, even toward targets defined in other people, and potentially during naturalistic experience. What remains untested is whether neurofeedback can guide a clinical population toward a clinically predictive neural target during a multimodal, socially rich, cue-laden movie that approximates the contexts in which recovery unfolds - and whether that learning is accompanied by meaningful improvement in clinical outcomes.

Implementing naturalistic movies and leveraging the evolving process of recovery in the design of rt-fMRI-NF paradigms could help close these gaps. Movies are increasingly recognized as useful tools for psychiatric neuroscience because they more effectively engage higher-order cognitive, affective and social processes than static picture cues, while maintaining a controlled yet ecologically valid format^29,30^. Moreover, they evoke temporally extended brain responses, making it possible to isolate neural dynamics shared within a group during rich, real-world-like experiences^31–33^. These rich dynamics can be leveraged for peer-guided learning, as the relevant target signal should not only be recovery-related, but also sufficiently shared across people to be communicated and reconstructed. In addiction, a drug-themed movie can establish a sustained drug context in which drug cues, alternative reinforcers, and emotionally charged social interactions unfold within the same narrative stream, thereby more closely approximating the environments in which recovery must be maintained in the real world. In our recent work, individuals with opioid use disorder (iOUD) showed shared drug-biased dynamics in the orbitofrontal cortex (OFC) during a drug-themed movie, *Trainspotting,* which were amenable to treatment and tracked craving reductions^34^. These findings suggest an altered valuation landscape in addiction and that brain dynamics from a drug-related movie can define a recovery-relevant neural workspace. Our hypothesis was that the shared component of these brain dynamics may provide a tractable substrate for peer-guided learning during treatment.

Here we introduce peer-guided movie neurofeedback (PG-MovieNF), an inter-individual rt-fMRI-NF framework in which treatment-related neural dynamics derived from individuals later in recovery are used as the learning target for individuals earlier in recovery. We first used longitudinal fMRI data from inpatient iOUD who viewed the same *Trainspotting* segment before and after 8 weeks of adjunctive treatment to derive a treatment axis capturing shared recovery-related change in movie-evoked brain dynamics. A multivariate signal from dorsolateral PFC (dlPFC) and dorsomedial PFC (dmPFC) regions of interest (ROI) was selected in this initial cohort based on hypothesized sensitivity to cognitive reappraisal strategies^35–37^ and cross-validated classification of pre- versus post-treatment responses. We then trained a separate cohort of iOUD to increase alignment with this peer-derived dlPFC/dmPFC signal during repeated viewings of the same movie, with feedback delivered at selected scene breaks throughout the movie. We asked whether participants could progressively align their brain dynamics with the peer-derived treatment signal, and given our prior work^34^, whether such alignment would engage shared OFC dynamics, decreasing craving and improving affect. Thus, we treated dlPFC/dmPFC dynamics as a trainable reappraisal and control-related entry point, and OFC dynamics as a downstream readout identified in our prior work. Real PG-MovieNF was compared with yoked sham feedback, whereby each participant viewed the feedback scores from an abstinence-matched participant from the real group, controlling for non-contingent feedback and repeated movie viewing while removing the coupling between the feedback and one’s own brain activity. Although tested here in iOUD, PG-MovieNF is intended as a general framework for using peer-derived neural dynamics to guide learning in distributed brain systems, with potential relevance for recovery in psychopathology and more broadly for learning adaptive brain states in naturalistic contexts.

## Results

### Identification of recovery-related target dynamics in a peer reference cohort

The first stage of PG-MovieNF is to identify treatment-related brain dynamics during a drug-themed movie that are shared by a group of peers and could be used as the neurofeedback target. For this target-identification stage, a peer reference sample of 37 inpatient iOUD watched the same 17-minute segment of the Oscar-nominated movie *Trainspotting* during fMRI, both before and after 8 weeks of adjunctive group therapy treatment (Figure 1A; see Table S1 for sample details; see Kronberg et al.^34^ for additional details). Our goal was to derive a spatial pattern of voxel weights (the *treatment transform*) that, when multiplied with the fMRI time series, results in a scalar time series with dynamics that best distinguish pre- from post-treatment (we refer to this scalar time series as a trajectory in *treatment space*; Figure 1B). Newly recruited peers can then be guided to adjust their brain dynamics via PG-MovieNF away from a pre-treatment peer template and towards a post-treatment peer template in this treatment space (Figure 1D top).

**Figure 1.**
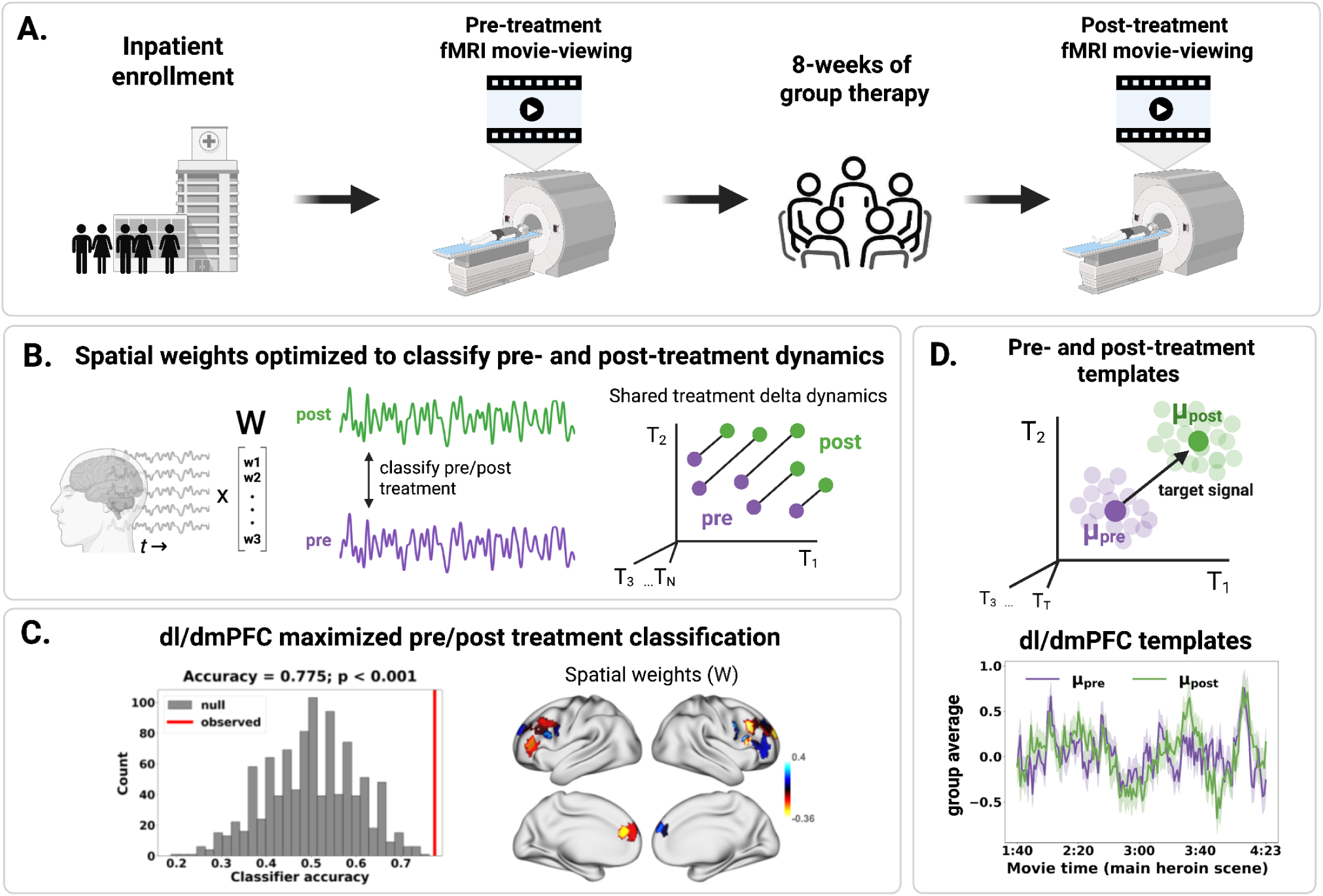
Target identification pipeline for deriving a shared, recovery-related signal for PG-MovieNF. **A.** Study Design. Thirty-seven iOUD watched the same 17-minute segment of the movie *Trainspotting* during fMRI both before and after 8 weeks of adjunctive group therapy, yielding paired pre- and post-treatment responses per subject. **B.** Deriving the treatment transform. **Left.** Regional time series are projected through a set of spatial weights (treatment transform, *W*) to yield a one-dimensional signal in ‘treatment space’. **Middle.** The goal, identify a treatment transform, *W*, with treatment-space dynamics that can be most accurately classified into pre- (purple) and post-treatment (green) sessions. **Right.** Visualization of treatment space dynamics, where each dimension corresponds to a TR during the movie (T_1_…T_T_), and each point corresponds to the full treatment space dynamics for a given session. Lines between paired points correspond to the treatment delta dynamics for a given subject. DeltaCorrCA maximizes the ISC of the treatment delta dynamics, which correspond to long parallel lines, enabling classification of the dynamics into pre and post treatment. **C.** Neural target. **Left.** The dl/dmPFC set of regions was selected as it maximized accuracy of classifying pre- vs. post-treatment sessions on held-out subjects (accuracy = 77.5%, p < 0.001). **Right.** Refitting on all 37 subjects yields the final transform *W* (visualized on the cortical surface) as well as the group-mean pre- and post-treatment template dynamics (*μ*_pre_ and *μ*_post_), visualized in **D.** Target signal. **Top.** PG-MovieNF uses a target that trains participants to make their treatment space dynamics move away from *μ*_pre_ and towards *μ*_post_. **Bottom.** The group-mean dl/dmPFC pre- and post-treatment templates, expanded during a main use heroin scene; shading denotes the confidence interval.

Our method for learning this treatment transform (see Methods section *Deriving the target signal*) involved finding a linear transformation of the fMRI time series that maximizes the inter-subject correlation (ISC) of the treatment delta signals (post-minus-pre treatment time series) (Figure 1B, right); intuitively, our goal here was to transform the data in a way that highlights differences in the post- and pre-treatment time series that are reliable across participants in the training sample, and therefore are likely to also be present in held-out participants. Correlated components analysis^38^ (CorrCA) is a principled method for identifying linear transformations that maximize ISC. We therefore applied CorrCA to the within-subject ROI-level treatment delta signals (deltaCorrCA) to identify a treatment transform. We tested several combinations of parameters (e.g. regularization and preprocessing choices) and pre-selected sets of ROIs, evaluated via classification accuracy of pre/post treatment labels with ten-fold cross validation stratified by subject (See supplementary section *Candidate Models Evaluated During Target Identification* and Table S2 for details).

The model that best distinguished pre from post-treatment included ROIs primarily from the bilateral dlPFC and some dmPFC (Brodmann areas 9 and 46), which achieved a mean classification accuracy on held out subjects of 77.5% (p<0.001; Figure 1C). After identifying this set of ROIs and parameters, the deltaCorrCA model was refit with all 37 peer reference iOUD, yielding a treatment transform (W) as well as pre-treatment and post-treatment template dynamics in the treatment space (*μ*_pre_ and *μ*_post_). The difference between these templates serves as the *target signal* for PG-MovieNF, teaching an independent cohort to move away from the pre-treatment and towards the post-treatment template dynamics in treatment space (Figure 1D).

### PG-MovieNF Task

An independent sample of 20 iOUD (10 real feedback, 10 yoked sham) completed up to eight runs of a novel PG-MovieNF task, during which they viewed the same segment of Trainspotting used in the target identification phase while receiving intermittent feedback between natural scene breaks. Each run contained 22 predefined feedback “stations”—temporally extended analysis windows in which the peer-derived target changed approximately monotonically—distributed across eight scored scenes. Stations within a scene were concatenated to generate one feedback score at the following scene break (Figure 2; see Methods). For the real group, each feedback score reflected the real-time similarity of their transformed signal to the treatment-related target signal; for the sham group, each participant viewed the feedback scores from a matched counterpart in the real group.

**Figure 2.**
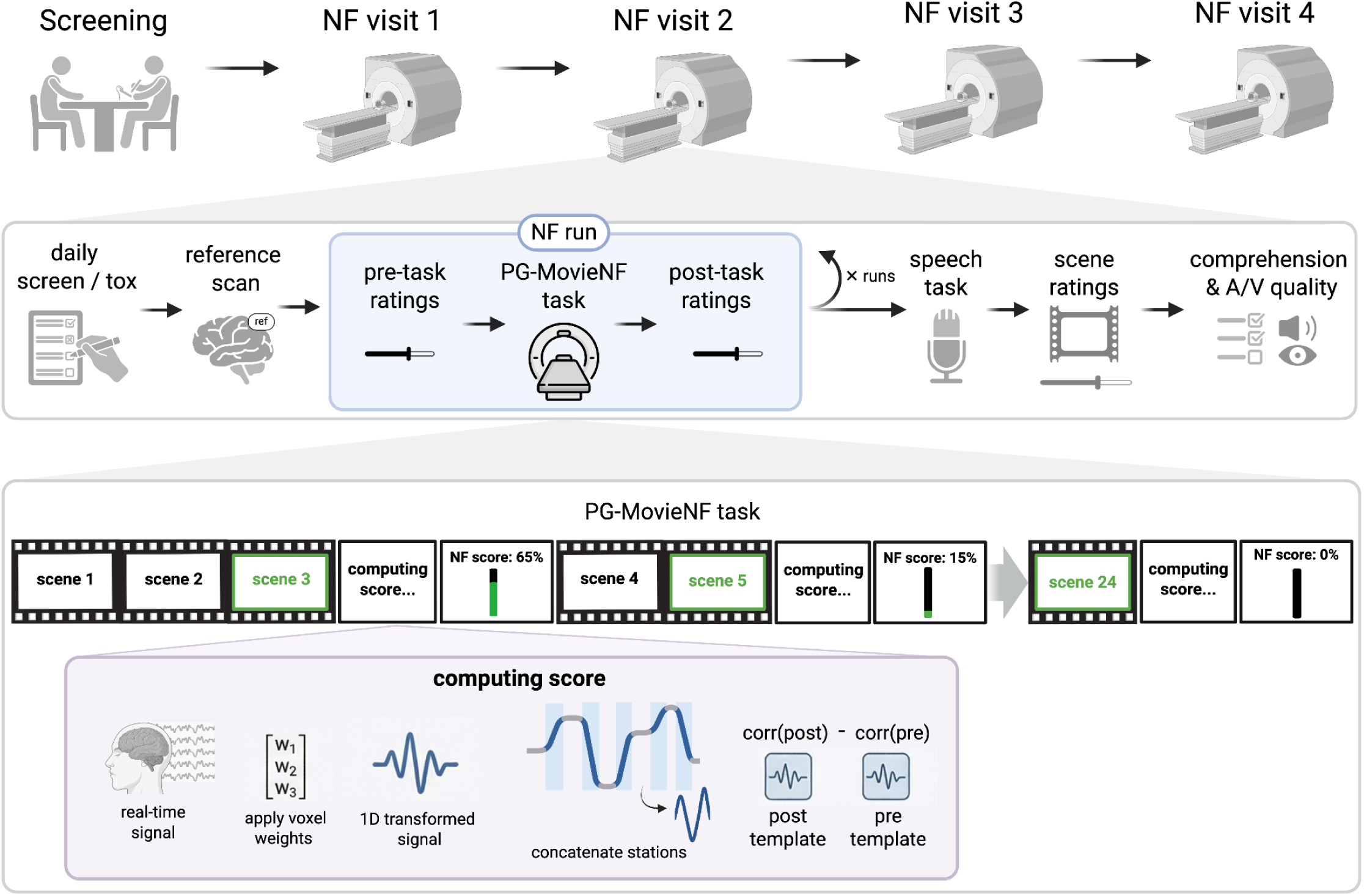
Study design and PG-MovieNF task schematic. **Top.** Study timeline: participants completed a screening visit followed by up to four neurofeedback visits. **Middle.** Structure of a neurofeedback visit: daily screen and urine toxicology, a reference scan for mask registration, one or more PG-MovieNF runs (each flanked by pre- and post-task ratings), a post-NF speech task, and a post-movie questionnaire assessing scene-level craving, arousal, and valence ratings as well as comprehension and audiovisual (A/V) quality. **Bottom.** PG-MovieNF task schematic: twenty four movie scenes are presented, each corresponding to a distinct narrative segment. There are a total of eight scored scenes, indicated by green borders and text. After each scored scene, a ‘computing score…’ display appears during which the real-time analysis pipeline applies voxel weights to the real-time fMRI signal to produce a transformed one-dimensional signal. Next, ‘stations’ (indicated in blue) are extracted – these are segments of the signal that were monotonically increasing or decreasing in the peer reference sample, likely reflecting a cognitive operation that can be manipulated (see Methods). These stations are then concatenated, and compared against pre- and post- treatment target templates to generate a scene-level neurofeedback score. After the score is finished computing, it is displayed to the participant before the task advances to the next scene (after a ‘continuing movie…’ display, which is not visualized).

### Testing PG-MovieNF efficacy

To test whether PG-MovieNF translated into clinical treatment outcomes, our primary outcome measure was scene-induced craving, collected at the end of each visit outside the scanner in response to short movie clips^34^. Affective state ratings^39^ collected before run 1 and after run 8 (pre and post PG-MovieNF overall) served as a secondary outcome measure. We hypothesized that the real group would exhibit an increase in neurofeedback scores over runs (reflecting learning of the target dynamics), reduced craving, improved affect, and significant coupling between one’s alignment with the target signal and shared OFC dynamics; these effects were expected both within the real group alone and compared to sham. We further hypothesized that, across participants, neurofeedback learning would correlate with craving and affect changes, while OFC coupling would correlate with craving changes. We treated these hypotheses as a family of statistical tests for the efficacy of PG-MovieNF, correcting for the false discovery rate (FDR) over all of them^40^. Neurofeedback learning after run 4 may have been disrupted due to reduced task engagement and real-time processing delays (Supplementary Fig. S1; see Supplemental section *Late-run task disruption motivates testing runs 1–4*). We therefore included both runs 1-4 (when available) and runs 1-8 in the main FDR-corrected family of tests. Given our strong directional hypotheses for each effect, we use one-sided p-values for this entire family of tests. We ran several analyses to test for baseline group differences in these measures, which are excluded from the main FDR correction and use two-sided p-values, as they solely aim to test for group equivalence as a control measure.

### Neurofeedback learning and associated improvements in craving and affect

To assess neurofeedback learning, we fit a linear mixed effects model predicting feedback scores from run, group (real, sham), and their interaction, with random slopes and random intercepts for subjects and real-sham pairs (see Methods). For runs 1-4, the real group showed a significant linear increase in neurofeedback performance across runs (Figure 3A, top; β = 0.37, t(18) = 2.34, one-sided p = .016, d = 1.10, FDR q = .031), whereas the sham group did not (β = 0.08, t(18) = 0.49, one-sided p = .314, d = 0.23). The group × run interaction was in the predicted direction (real > sham; β = −0.15, t(18) = −1.30, one-sided p = .105, d = −0.61, FDR q = 0.145), although it did not reach significance. Groups did not differ at baseline (β = 0.22, t(16) = 1.00, two-sided p = .332, d = .49). When all 8 runs were included, the learning effect in the real group was attenuated (β = −0.06, one-sided p = .721, FDR q = 0.721), as was the group × run interaction (β = −0.07, one-sided p = .191, FDR q = 0.229); the sham group also did not increase across this run window (β = −0.193, one-sided p = .963).

**Figure 3.**
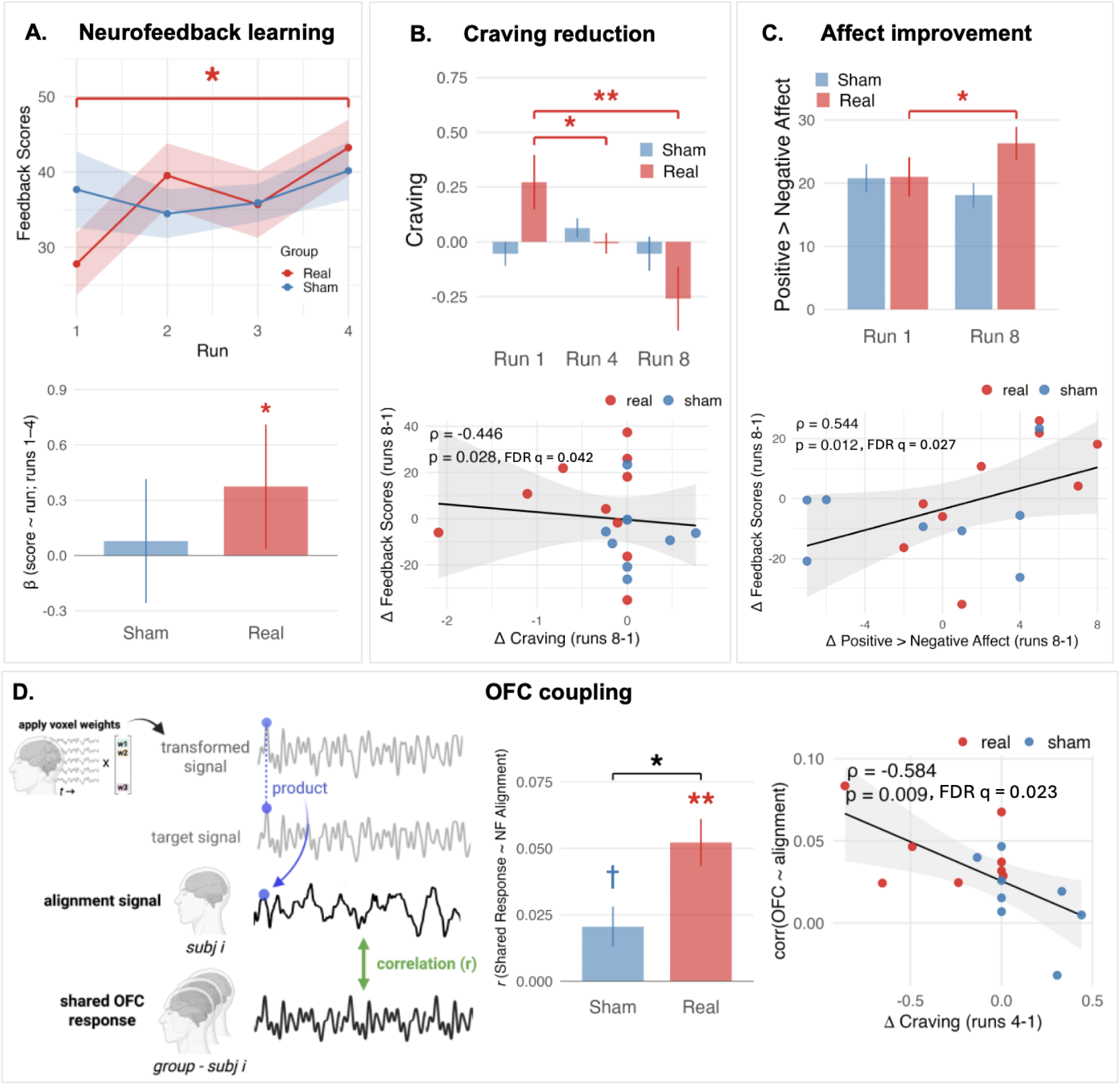
Neurofeedback learning, improvements in craving and affect, and OFC coupling. **A.** Neurofeedback performance. **Top:** Mean neurofeedback scores for the real and sham groups across the first four runs. **Bottom:** Standardized β for the effect of run on feedback score. Error bars indicate 95% confidence intervals. **B.** Scene-induced craving. **Top:** Craving scores at runs 1, 4, and 8 by group. Four subjects were excluded from the run 1–4 comparison because runs 1 and 4 fell within a single session, precluding delta computation. **Bottom:** Spearman correlation between change in neurofeedback scores and change in craving (run 1-8; n=19 as one subject was missing run 8 feedback scores). **C.** Positive > negative affective ratings. **Top:** Ratings at runs 1 and 8 by group. **Bottom:** Spearman correlation between change in neurofeedback score and change in positive > negative affective ratings [run 1-8; n = 17 after excluding one outlier (± 3 SD), and one subject missing run 8 feedback scores and another missing run 1 affective ratings]. **D.** OFC coupling with alignment dynamics. **Left:** Schematic of the analysis pipeline. For each scan, voxel weights were applied to the dl/dmPFC BOLD time series to yield a 1D treatment space signal. The elementwise product of this treatment space signal with the target signal produced an ‘alignment’ signal, in which positive values index moments when treatment space dynamics are aligned with the target dynamics. Similar to an inter-subject functional connectivity approach, concatenated alignment signals across all training runs were then correlated (via Pearson correlations) with a leave-one-out group shared OFC response estimated using a Shared Response Model. **Middle:** Coupling between each participant’s alignment dynamics and the shared OFC response during feedback stations. One-sample t-tests on the correlation coefficients revealed that both groups showed significant coupling (real: t(9)=5.89, one-sided p<.001, FDR q = .002; sham: t(9)=2.72, one-sided p=.012), with stronger coupling in the real than the sham group (t(18)=2.72, one-sided p=.007, FDR q = .023). **Right:** Across participants, stronger shared OFC-alignment coupling was associated with larger reductions in scene-induced craving from run 1 to run 4 (⍴ = −0.584, one-sided p = .009, FDR q = .023). Bar plots visualize means ± SE (except in **A**). For correlations, shaded bands show 95% confidence intervals around the regression line, and ρ reflects the full sample Spearman correlation. Colored brackets denote significant within-group comparisons; black brackets denote significant between-group comparisons; colored asterisks above individual bars denote significant one-sample tests. Given strong directional hypotheses for these effects, one-sided tests were used throughout. Asterisks denote the Benjamini–Hochberg FDR q-value for tests included in the correction; *<.05; **<.01; ***<.001. Dagger denotes uncorrected p-value for tests not included in the correction; †<.05.

The real group showed significant reductions in craving across runs 1-4 (β = −0.59, t(14) = −2.76, one-sided p = .008, d = −1.95, FDR q = .023), and runs 1-8 (β = −0.66, t(38) = −3.93, one-sided p < .001, d = −2.49, FDR q = .002), whereas the sham group did not (runs 1-4: β = 0.25, t(14) = 1.17, one-sided p = .868, d = 0.82; runs 1-8: β = 0.06, t(38) = 0.38, one-sided p = .647, d = 0.24; Figure 3B, top). The group × run interactions confirmed steeper declines in craving in the real group in both run windows (1-4: β = 0.84, t(14) = 2.78, one-sided p = .007, d = 1.96, FDR q = .023; 1-8: β = 0.72, t(56) = 3.05, one-sided p = .002, d = 1.93, FDR q = .010). While the real group showed higher craving than the sham group after run 1 (runs 1-4 sample: β = −0.70, t(28) = −3.08, two-sided p = .005, d = −1.54; runs 1-8: β = −0.46, t(56) = −2.54, two-sided p = .014, d = −1.13), we confirmed that the pre-neurofeedback craving measures did not differ between groups (two-sided ps ≥ .197), nor did controlling for them affect the group × run interaction effects (one-sided ps < .007; see Supplemental section *Controlling for baseline craving in the craving reduction models*). Larger improvements in feedback scores tracked larger reductions in craving across runs 1-8 (n=19; ρ = −.446, one-sided p = .028, FDR q = 0.042), an effect not evident at runs 1-4 (n=16; ρ = −.101, one-sided p = .355, FDR q = 0.399; Figure 3B, bottom).

Affective state also improved selectively in the real group, which showed an increase toward positive relative to negative affect from run 1-8 (β = 0.33, t(17) = 2.16, one-sided p = .023, d = 1.36, FDR q = .037), while the sham group did not (β = −0.16, t(18)= −1.00, one-sided p = .834, d = −0.63; Figure 3C, top). The group × run interaction confirmed a greater shift in the real group than the sham (β = −0.49, t(18) = −2.22, one-sided p = .020, d = −1.40, FDR q = 0.036; Figure 3C, top). Baseline affect did not differ between groups (β = −0.02, t(20) = −0.09, two-sided p = .929, d = −0.04). Greater increases in feedback scores tracked greater improvement in affect (n = 17; ρ = .544, one-sided p=.012, FDR q = 0.027; Figure 3C, bottom).

### Shared orbitofrontal dynamics track alignment with treatment-related dynamics

We hypothesized that successful target alignment would be coordinated with broader neural processes relevant to treatment response. The OFC was a strong candidate because our prior work in this population identified treatment-sensitive shared OFC dynamics, with OFC synchrony tracking reductions in craving^34^. These findings positioned shared OFC dynamics as a candidate readout of how the drug narrative was represented over the course of training, and we hypothesized that this readout would be coordinated with successful target alignment. Such coordination should be evident as moment-to-moment covariation between target alignment and shared OFC dynamics.

To test this hypothesis, we computed a time-resolved alignment signal as the TR-wise product of each participant’s treatment-space signal and the target signal, with positive values indicating momentary alignment (Figure 3D). Because our hypothesis concerned coordination between this alignment signal and OFC dynamics that are shared across subjects, we used an inter-subject functional connectivity (ISFC) approach^32^, where each participant’s alignment signal during feedback intervals was correlated with the shared OFC response estimated from the other participants in the same condition using a Shared Response Model (SRM; see Methods). This approach emphasizes stimulus-locked OFC dynamics common across viewers while reducing noise and idiosyncratic contributions.

The real group exhibited a significant coupling of shared OFC responses and alignment dynamics (M = 0.052, SD = 0.028, *t*(9) = 5.89, one-sided *p* < 0.001, FDR q = .002); to a lesser extent, the sham group exhibited significant coupling as well (M = 0.021, SD = 0.024, t(9) = 2.72, one-sided *p* = 0.012). This OFC-alignment coupling was significantly stronger in the real than the sham group (Figure 3D, middle; *t*(18) = 2.72, one-sided *p* = 0.007, FDR q = 0.023). These effects were specific to runs 1-8 and feedback stations only, as they were not observed across the whole-movie for runs 1-8 (*p* ≥ 0.087), nor when restricted to feedback stations in runs 1-4 (between groups: t(18) = 0.93, one-sided *p* = 0.183, FDR q = 0.229; see Methods). ISC of the alignment signal alone did not differ between groups, whereas ISC of the OFC signal alone was higher in the real group (See Supplement *Synchronization in OFC dynamics and alignment dynamics*). Thus, stronger real-group OFC–alignment ISFC coupling may partly reflect a more reliably shared OFC response in the real group.

We then examined whether the degree of OFC-alignment coupling predicted greater reductions in craving, and found that participants with stronger coupling showed greater decreases in scene-induced craving from runs 1-4 (n=16, ⍴ = −0.584, one-sided p = 0.009, FDR q = 0.023; Figure 3D, right; see Methods for subsampling details). This association was not significant when extended to run 1-8 craving reduction (n=20, ⍴ = −0.009, one-sided p = 0.485, FDR q = 0.514).

As an exploratory analysis, we further tested the directionality of the OFC-alignment coupling. We fit bivariate vector autoregressive models and tested Granger causality in both directions (OFC → alignment, alignment → OFC). Alignment dynamics significantly preceded shared OFC responses in the real group (*t*(9) = 3.02, two-sided *p* = 0.014) but not in the sham group (two-sided *p* = 0.810), and this difference was significant between groups (*t*(18) = 2.15, two-sided *p* = 0.045). The reverse direction was not significant in either group or between groups (two-sided *p* ≥ 0.128).

Beyond the OFC, convergent brain dynamics, as measured by ISCs across the whole movie, emerged across the prefrontal cortex, whereby the real group showed increasing ISCs in salience/ventral-attention frontal opercular regions, whereas the sham group showed decreasing ISCs (Figure S2; see Supplemental section *Real PG-MovieNF increases prefrontal inter-subject synchronization*). Together, these findings indicate that the real PG-MovieNF group exhibited convergence toward a more shared mode of prefrontal processing of the drug-related narrative, whereas non-contingent sham feedback was associated with progressive desynchronization.

## Discussion

### Main findings and conceptual advance

PG-MovieNF was designed to test the largely unexplored possibility that recovery-relevant brain dynamics identified in one group of peers can guide learning in another. Using a longitudinal dataset in which heroin-primary iOUD watched a drug-themed movie during fMRI before and after treatment, we identified a peer-derived, recovery-related target signal; namely a shared dl/dmPFC-weighted signal that maximally distinguished pre- from post-treatment responses. We then tested whether an independent cohort of iOUD could learn to align their brain activity with this target signal during repeated viewing of the same movie. Participants in the real group received feedback based on their real-time alignment with the post-treatment template relative to the pre-treatment template. Participants in the yoked sham condition received feedback that was identical to their yoked pair in the real group, but unrelated to their own brain activity.

Real PG-MovieNF showed a clear and convergent pattern of improvement across the study’s central outcomes, including increases in target alignment over runs 1–4, reduced scene-induced craving, and improved positive relative to negative affect, increased prefrontal ISC, and strengthened coupling between dl/dmPFC treatment-signal alignment and shared OFC responses, which was predictive of movie-induced craving reduction. Scene-induced craving provides the most direct test of transfer from PG-MovieNF to clinical outcome because it measures how participants responded to the same drug-relevant narrative context used during training. Given that the affective ratings were not elicited in response to the movie, their improvement suggests that PG-MovieNF may have shifted participants’ overall emotional state. Importantly, the degree of symptom improvement was directly linked to the degree of neurofeedback learning. These findings establish proof of concept for a socially scaffolded neurofeedback framework, here tested in addiction. This approach marks a shift away from neurofeedback training of an individual’s own single-region brain activity in response to static images, and toward training to align distributed brain dynamics with a target signal derived from peers, embedded within a shared naturalistic recovery-relevant context.

### Relationship to prior neurofeedback work

Rt-fMRI-NF has previously shown some promise for modulating addiction-related brain activity: Recent reviews conclude that rt-fMRI-NF can alter craving-related brain function and subjective craving^21,41^, but most studies have focused on nicotine, alcohol, and cocaine rather than opioids^21^. Further, while some recent addiction neurofeedback work has begun to move toward more distributed targets^42–45^, the field has been dominated by the use of univariate target signals in combination with static drug cues (mostly pictures), e.g., using individualized cue-reactivity regions or predefined craving-control ROIs^46–51^.

Beyond addiction, the rt-fMRI-NF literature more broadly has developed sophisticated multivariate feedback signals, e.g., based on functional connectivity^52^, multivariate activity patterns^23,25,27,53^, and targets defined from independent participants or cohorts^26^. Mennen et al.,^28^ whose approach inspired much of the design for the present study, extended these approaches to a naturalistic audio narrative stimulus. However, no studies that we are aware of have used rt-fMRI-NF during an extended movie stimulus. Mennen et al.^28^ trained a classifier to predict participants’ interpretations of an ambiguous narrative in a pre-training cohort. They then used the pre-trained classifier to provide feedback and guide narrative interpretations in an independent cohort using rt-fMRI-NF. Here we extended this general design to an engaging movie that is tailored to a psychiatric population and a target signal that is optimized with a longitudinal dataset to maximize sensitivity to treatment effects. PG-MovieNF therefore represents a unique intersection of methodologies as the first study using longitudinal data to define shared, multivariate, recovery-related neural dynamics during a disorder-relevant movie and then training a group of peers with the same psychiatric disorder to reproduce these dynamics via neurofeedback.

### Potential mechanisms

The dl/dmPFC-weighted target builds directly on the impaired response inhibition and salience attribution (iRISA) framework, which emphasizes an imbalance between exaggerated salience attribution to drug-related cues and impaired recruitment of prefrontal systems supporting inhibitory control and related functions (e.g., self-monitoring/regulation, goal maintenance, and adaptive valuation)^11,12,54,55^. In iOUD specifically, we recently reported that anterior PFC engagement during inhibitory control (including dlPFC) was reduced at baseline compared to healthy controls, and increased after inpatient treatment^56,57^. A complementary drug cue-reactivity study showed that drug cue reappraisal recruited the dlPFC, with stronger dlPFC engagement during reappraisal associated with lower cue-induced craving^58^. While these findings support the use of dl/dmPFC as a clinically meaningful control-related search space, only a small number of addiction rt-fMRI-NF studies have targeted dl/dmPFC ROIs. These include a dmPFC arm in smokers^46^, individualized ROI approaches in which the dlPFC was one possible target region^48,59^, and a broader multi-ROI approach that included the dlPFC as one component of the feedback signal^44^. Thus our study is unique in explicitly targeting the dl/dmPFC in addiction, with Brodmann areas 9 and 46 constraining target discovery to an anatomically interpretable territory. This ROI choice was further motivated by dl/dmPFC sensitivity to longitudinal treatment effects in our peer reference sample, unlike previous studies that derived neurofeedback targets based on drug cue-reactivity at a single time point.

The OFC findings extend our prior work implicating synchronized OFC activity as a naturalistic marker of drug-biased salience, craving, and recovery^34^. Here, alignment with the peer-derived dl/dmPFC recovery signal was more strongly coupled to shared OFC responses during real PG-MovieNF, and this coupling correlated with greater craving reduction across participants. This finding suggests that participants were not only changing activity in the feedback target, but engaging a control state that altered how the drug narrative was valued as it unfolded. This account fits contemporary theories of OFC function as representing outcome value and cognitive maps of motivationally relevant states, allowing behavior to update when goals, needs, or contexts change^60,61^. In addiction, recovery may require updating the expected value of drug-related outcomes relative to alternative reinforcers, future consequences, and current treatment goals. PG-MovieNF may therefore act at a control–valuation interface: dl/dmPFC alignment may help guide OFC representations of the drug context toward a recovery-consistent map. The finding that alignment preceded shared OFC responses in the real group is consistent with this account, although it should be interpreted as correlational mechanistic support rather than evidence of causation. This account is consistent with primate work showing that dlPFC and OFC contribute distinct signals during reward-guided choice, with dlPFC activity tracking whether task-relevant performance goals are met and OFC activity tied more closely to reward as it is encountered^62^ (see also Wallis & Miller^63^).

Although OFC responses were treatment-sensitive in our previous study^34^, OFC-based models were not among the strongest candidates for deriving the neurofeedback target signal (see Supplementary Table S2). This apparent discrepancy likely reflects different analytic objectives. The event-based reverse-correlation analysis from the prior study was done independently in each parcel and tested whether extreme shared responses, including both peaks and troughs, preferentially followed drug content in iOUD relative to healthy controls. Target identification here instead required a continuous, multiregion trajectory that generalized across individuals and distinguished pre- from post-treatment scans in iOUD over the full movie. The OFC–alignment analysis in the present study further asked whether a single shared OFC trajectory covaried with moment-to-moment target alignment. These three analyses therefore probe complementary properties of OFC processing—event-specific sensitivity to drug content, whole-movie treatment-stage discriminability, and moment-to-moment coordination with target alignment. Because significant drug-content selectivity in the prior study^34^ and target-alignment coupling here were observed within selected movie segments, treatment-sensitive OFC processing may be context-dependent rather than expressed as a stable whole-movie recovery signal. This interpretation is also consistent with OFC–alignment coupling observed during intervals used as feedback stations but not across the full movie.

Real PG-MovieNF also increased prefrontal ISC across participants, whereas yoked sham feedback did not, pointing to a complementary mechanism in line with the inter-individual nature of the target signal. Increased ISC in the real group suggests that participants receiving contingent feedback converged toward a more common mode of processing the movie. This synchrony was not related to the feedback score itself, indicating that PG-MovieNF may have promoted broader convergence of prefrontal systems involved in control and regulation during the drug-related narrative.

### Broader implications

Although developed for addiction recovery, PG-MovieNF defines a more general strategy for training adaptive brain dynamics during clinically meaningful experiences. The framework has three core components: a naturalistic context that evokes symptom-relevant processing, a reference trajectory derived from peers further along in the learning process, and real-time feedback that trains others to move toward that trajectory. In addiction, the relevant context is a drug-related narrative and the target is a peer-derived recovery signal. In other conditions, the same logic could be adapted to different symptom-relevant contexts, including trauma, social threat, uncertainty, compulsive concerns, or body/food related cues. The goal would not be to impose a generic “healthy” brain state, but to identify adaptive trajectories that emerge during meaningful clinical change and train patients to approach those trajectories while immersed in the contexts where symptoms arise.

Beyond its clinical applications, PG-MovieNF offers a framework for testing the causal and behavioral impact of the shared brain dynamics identified in the growing body of movie fMRI data and inter-subject analyses. Movie fMRI and inter-subject analyses can identify shared neural trajectories associated with symptoms, traits, and behaviors, but they remain largely correlational. The PG-MovieNF approach offers a way to perturb a shared neural state space and causally test whether eliciting specific shared dynamics also produces hypothesized downstream neural or behavioral effects.

### Limitations and future directions

These findings should be interpreted in light of several limitations. First, this was a proof-of-concept study with a modest sample size, so the results should be understood as evidence of feasibility and mechanistic promise rather than clinical efficacy. Groups were also enrolled sequentially rather than randomized, the sham group having been recruited after feasibility was established in the real group, so cohort effects cannot be excluded; the groups were yoked on abstinence duration and did not differ at baseline on demographic, clinical, substance use, or pre-neurofeedback craving measures. The yoked sham condition controlled for movie exposure, feedback timing, visible score structure, and the experience of receiving plausible feedback, but future studies should also compare PG-MovieNF against additional active controls that test the specificity of the target signal in conferring clinical value, e.g., active controls using alternative movie-responsive brain signals, orthogonal dl/dmPFC dimensions, and strategy-only training (i.e. no neurofeedback).

The current protocol also leaves open questions about neurofeedback dose and implementation. The clearest learning effects were observed early in training, particularly across runs 1–4, whereas later runs were affected by reduced engagement and technical limitations. This pattern may reflect both task burden and implementation constraints: the protocol may have exceeded the optimal training dose, and later runs were more vulnerable to disruptions that reduced feedback reliability. Future versions could therefore emphasize a shorter, more reliable, and more interpretable training window. Nevertheless, results that were unique to all 8 runs (e.g., correlation between neurofeedback scores and craving) suggest extended training may be needed to reveal the full scope of the effects. More generally, several effects and their clinical correlates fell in different run windows: learning was significant across runs 1–4 but tracked craving reduction only across runs 1–8, whereas OFC-alignment coupling differed between groups across runs 1–8 but tracked craving reduction only across runs 1–4. Both run windows were included in the FDR-corrected family of tests; resolving this dissociation will require a design with a prespecified training length.

The neural target and mechanistic analyses also require cautious interpretation. The dl/dmPFC signal was selected because it was empirically effective and theoretically grounded, but Brodmann areas 9 and 46 provide a coarse anatomical search space that includes functionally heterogeneous prefrontal territories. The target should therefore be interpreted as a useful control-related trajectory, not as evidence that recovery localizes specifically to BA9/46. Similarly, the OFC coupling and directionality analyses are mechanistically informative but correlational. The finding that target alignment preceded shared OFC responses is consistent with a control-to-valuation account, but future studies with causal manipulations specifically designed to test this mechanism are needed.

Finally, the clinical reach of PG-MovieNF warrants further research. The present study tested individuals with heroin-primary OUD in inpatient treatment, where medications for OUD and motivation to abstain may shape both movie responses and neurofeedback learning. It is therefore unknown whether the same target, stimulus, or training structure will generalize to outpatient populations, other substance use disorders, or individuals in different stages of recovery. Moreover, while the scene-induced craving and affect outcomes are clinically meaningful, future work could test whether PG-MovieNF produces long-lasting changes in craving, treatment retention, and drug use patterns, or other objective behavioral markers^64^. Given the limited immediate scalability of fMRI, more deployable approaches using EEG, fNIRS, eye gaze, or stimulation-based interventions could be considered for related target discovery and mechanism validation.

## Methods

### Peer Reference Sample and Dataset

The peer reference dataset consisted of functional magnetic resonance imaging (fMRI) blood oxygenation level dependent (BOLD) activity collected from 37 iOUD (see Table S1 for sample characteristics) while they watched the first 17 minutes of the movie ‘Trainspotting’ twice: once pre-group-therapy (177.6 ± 128.1 days into enrollment in a residential inpatient treatment facility, receiving standard of care) and once post-group-therapy (100 ± 34 days later); this therapy consisted of an 8-week randomized adjunctive group treatment as part of an associated clinical trial (NCT04112186; for more details, see Kronberg et al. and Huang et al.^34,65^). At baseline, all iOUD were abstinent and stabilized on medication for opioid use disorder (MOUD; confirmed via urine toxicology) initiated upon treatment enrollment. All met criteria for opioid use disorder (assessed via the Mini-International Neuropsychiatric Interview, 7th ed.) with heroin as the primary reason for treatment (assessed via a modified Addiction Severity Interview, 5th ed.). For further details on this dataset, including anatomical, functional, and custom preprocessing steps applied to the neuroimaging data, see Kronberg et al^34^.

### Deriving the target signal

Our goal was to derive a one-dimensional movie-evoked signal that captured treatment-related dynamics shared across the peer reference cohort and could be applied to new participants during real-time neurofeedback. For subject *i* and session *j* ∈ pre,post, let 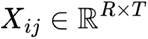 denote the processed BOLD time series from *R* regions of interest (ROIs) across *T* repetition times (TRs). We sought a spatial weight vector *W* ∈ ℝ^R^ that projected these multivariate responses onto a scalar time series,

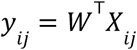

We refer to the one-dimensional coordinate defined by *W* as treatment space and to *y_ij_* as the treatment-space signal. The intended neurofeedback target was not a sustained increase or decrease in this signal. Rather, PG-MovieNF participants were trained to make its temporal dynamics more similar to a peer-derived post-treatment template and less similar to a pre-treatment template. We therefore aimed to select *W* such that pre and post treatment labels can be most accurately classified from the full dynamics of *y_ij_*. To identify treatment-related dynamics, we first computed each subject’s within-subject treatment-delta matrix,

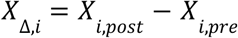

We then applied CorrCA to the treatment-delta matrices across subjects. CorrCA identifies a common spatial projection for which the resulting time series are maximally correlated across individuals, i.e. ISC is maximized^38^. Applied here, it finds *W* such that the projected treatment-delta signals,

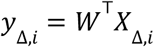

have maximal ISC. We refer to this application as deltaCorrCA. Conceptually, deltaCorrCA identifies a dimension along which post-minus-pre treatment changes in movie-evoked dynamics follow a similar temporal trajectory across the peer-reference cohort. The resulting component is therefore both treatment-related, because it is estimated from within-subject change, and shared across peers, because the change is optimized for reliability across individuals. In the supplement we provide a derivation showing that the deltaCorrCA approach is formally related to maximizing classification accuracy of pre and post-treatment labels with the template-matching classifier described below (see Supplemental section *Formal derivation for deltaCorrCA*).

We evaluated candidate models using ten-fold cross-validation with folds defined at the subject level, such that both sessions from a participant were assigned to the same fold. Within each fold, deltaCorrCA was fit using only the treatment-delta data from the training subjects. The resulting transform was then applied separately to their pre- and post-treatment sessions. Projected signals were z-scored across time and averaged across training subjects to produce a pre-treatment template, *μ*_pre_, and a post-treatment template, *μ*_post_. Each held-out session was projected using the training-fold transform and classified according to the template with which it had the higher Pearson correlation. We used template matching because the same quantities—similarity to the post-treatment template relative to the pre-treatment template—were subsequently used to generate neurofeedback.

Candidate models were defined by a priori ROI sets and two additional parameters: the CorrCA regularization value and whether an overall mean movie-evoked time course was removed before fitting the model. We tested regularization values of 0,0.1, 0.2, and 0.5. Model performance was quantified as mean held-out pre-versus-post classification accuracy across the ten folds; the complete model comparison is reported in Table S2. The highest-performing model used bilateral dorsolateral PFC and adjacent dorsomedial PFC ROIs defined by Brodmann areas 9 and 46, with a regularization value of 0.1 and mean subtraction. This model achieved a mean held-out classification accuracy of 0.775. For this selected model, the observed accuracy exceeded all accuracies obtained from 1,000 within-subject randomizations of the pre- and post-treatment labels (empirical *p*=.001).

After selecting the ROI set and model parameters, deltaCorrCA was fit using all 37 peer reference participants. This yielded the final treatment transform *W* and the group-average treatment-space templates *μ*_pre_ and *μ*_post_. Together, these quantities defined the peer-derived neurofeedback target: ongoing activity in the independent neurofeedback cohort was projected into treatment space using *W*, and feedback reflected movement toward *μ*_post_ relative to *μ*_pre_.

### Feedback stations

Because the treatment-related target defined above is a continuous template over the full movie, whereas neurofeedback was delivered intermittently between scenes, we next converted the continuous target into a set of discrete feedback stations. The goal of this step was to translate a complex continuous template into units that were interpretable for participants, compatible with slow hemodynamic responses, and minimally disruptive to movie immersion. Following Mennen et al.,^28^ we constrained station boundaries to natural scene breaks; we also selected contiguous movie intervals over which the target signal evolved approximately monotonically. This allowed each feedback score to summarize a temporally extended and directionally coherent segment of the target dynamics, more likely reflecting coherent cognitive and affective processing, rather than brief high-frequency fluctuations that are unlikely to be interpretable or controllable in real time. Stations were restricted to be at least 10 TRs long, monotonic when smoothed with a 10 TR moving average, and have mean pre/post treatment classifier accuracy greater than 0.5. This yielded 22 total stations throughout the 17 minute movie stimulus. Since feedback was delivered between natural scene breaks, multiple stations could fall within the same scene for feedback. In total, the 22 stations fell within 8 scenes. For scenes with multiple stations, the stations were first concatenated in time before deriving a single score to be displayed at the end of the scene.

### Neurofeedback sample

Ten iOUD were first recruited from the same medication assisted inpatient rehabilitation facility as the peer reference sample to receive real PG-MovieNF (real group). Following initial evidence of feasibility in the real group, an additional 10 iOUD were recruited to receive sham feedback (sham group). Each sham participant was yoked to a real group participant matched on abstinence duration (± 30 days) such that they viewed identical feedback, differing only in whether the feedback reflected the participant’s own real-time brain activity or those of their yoked counterpart. Similar to the peer reference sample, the iOUD in this sample were also abstinent (171 ± 92 days) and stabilized on MOUD (confirmed via urine toxicology) initiated upon treatment enrollment (162 ± 92 days prior to first neurofeedback visit). See the Supplement for further details regarding eligibility, diagnostic interviews, and the baseline assessments presented in Table 1. The real and sham groups did not differ on any demographic, clinical, or substance use characteristic (all *p*s > .015, Bonferroni-corrected for 20 comparisons; see Table 1).

**Table 1.** Neurofeedback sample characteristics by group. Continuous variables are presented as mean (standard deviation) and were compared using paired t-tests. Categorical variables are presented in counts; inferential statistics are not visualized as the small number of yoked pairs (N=10) produced sparse contingency tables, precluding valid paired categorical testing. Correcting for familywise error (α=.05/20=.0025), none of the variables exhibited significant between-group differences. All measures were collected at screening. ^a^Wilcoxon signed-rank tests were used, as assumptions of normality were violated.

|  | Real (n=10) | Sham (n=10) | sig. test |
| --- | --- | --- | --- |
| Age | 43.82 (10.82) | 40.90 (7.19) | 0.579 |
| Sex (Male/Female) | 9/1 | 10/0 |  |
| Race (White/Black/Other) | 6/2/2 | 5/2/3 |  |
| Ethnicity (Hispanic/Non-Hispanic) | 4/6 | 4/6 |  |
| Education (years) | 12.80 (1.93) | 10.90 (2.18) | 0.103 |
| Verbal IQ | 101.30 (10.69) | 93.60 (12.00) | 0.127 |
| Nonverbal IQ | 10.40 (1.17) | 11.00 (2.16) | 0.526 |
| Handedness (Right/Left) | 9/1 | 9/1 |  |
| Beck Depression Inventory (BDI) | 11.30 (8.18) | 9.00 (5.35) | 0.484 |
| Beck Anxiety Inventory (BAI) | 7.10 (6.90) | 3.60 (3.41) | 0.209 |
| Smoking Status (Current/Past/Never) | 10/0/0 | 8/1/1 |  |
| Fagerström Test for Nicotine Dependence (FTND) | 4.00 (1.56) | 3.50 (2.55) | 0.647 |
| Short Michigan Alcoholism Screening Test (SMAST) | 3.80 (3.16) | 3.40 (1.65) | 0.740 |
| Lifetime Heroin Use (years) | 16.90 (8.92) | 15.00 (6.41) | 0.623 |
| Heroin Abstinence (days) | 175.90 (94.29) | 165.30 (93.40) | 0.186 |
| Heroin Use Past Month (days) | 0.00 (0.00) | 0.10 (0.32) | 1.000<sup>a</sup> |
| Heroin Craving Questionnaire (HCQ) | 35.10 (6.79) | 37.70 (7.56) | 0.536 |
| Subjective Opiate Withdrawal Scale (SOWS) | 2.50 (4.35) | 0.80 (1.32) | 0.344<sup>a</sup> |
| Severity Of Dependence Scale (SDS) | 10.20 (3.33) | 12.90 (2.60) | 0.015 |
| Methadone Dosage (mg) | 109.00 (80.62) | 70.00 (70.18) | 0.224 |

The peer-reference and neurofeedback studies were approved by the Institutional Review Board of the Icahn School of Medicine at Mount Sinai. Participants provided informed consent and were compensated for study procedures.

### Neurofeedback task design

Having derived the peer-based target signal and feedback stations described above, we next implemented the neurofeedback task over repeated exposures to the same Trainspotting movie segment. We chose a total of eight runs to conservatively provide participants with sufficient opportunity to learn the task and to make it possible to detect learning-related change across training. This schedule was intended to reduce the risk that weak learning effects would be attributable simply to insufficient practice, with the majority of fMRI-NF studies in addiction relying on single-session designs, administering no more than two to three runs of neurofeedback^21^. The intended schedule comprised four visits on four consecutive days, with two runs per visit (Figure 2, top). When scheduling constraints prevented this nominal 4×2 structure, we prioritized completion of all eight runs on consecutive days, allowing some participants to complete fewer visits and up to four runs within a single visit. The specific placement of runs across sessions did not have a significant effect on neurofeedback scores when controlling for cumulative run number (see Supplemental section *Effects of deviations from the intended run structure*).

Before getting in the scanner on the first visit, all participants were informed of the overall study design (except the existence of a sham group) and goal of the study. Several suggestions were given for cognitive strategies to improve scores (see Supplement for more details), but participants were told to ultimately identify any strategy that they thought worked for them by paying attention to thoughts, feelings, or emotions and how they affect their score. Participants then completed a 5-minute practice run of the task with an unrelated non-drug movie to make sure they understood the task and were familiar with the format. On subsequent visits, participants were reminded of the task structure, suggestions for cognitive strategies, and were also prompted to recall their strategies from the previous visit.

Each run consisted of the same 17-min Trainspotting segment, containing eight feedback scenes which contained target stations (Figure 2, bottom). During any such feedback scenes containing one or more stations, a green border appeared around the screen for the full scene to indicate that the scene would contribute to the upcoming score. After each scored scene, the movie paused and a “computing score” screen was shown for 10 s. The feedback score was then displayed for 10 s as a green bar filled in proportion to the score, with the numeric value shown above the bar (e.g., “Neurofeedback score: 75%”). A “continuing movie” screen was then shown for 2 s before movie playback resumed.

### Real-time preprocessing and neurofeedback computation

PG-MovieNF was implemented using RT-Cloud^66^, installed locally on a dedicated analysis computer rather than deployed on a remote cloud server. The MRI scanner exported each reconstructed EPI volume as a DICOM file to a shared local network directory monitored by RT-Cloud, which converted incoming DICOMs into BIDS-compatible single-volume NIfTI images. A separate computer ran the PG-MovieNF task with custom PsychoPy code. The task and analysis computers communicated through the shared local network drive by writing and monitoring run-specific status and feedback-score text files. A public implementation of the real-time analysis and PsychoPy task is available online (see Code Availability).

At the beginning of each visit, participants completed a 1-min resting BOLD scan that provided the reference image for real-time processing. Resting volumes were motion corrected and averaged, and the resulting reference image was brain extracted using BET^67^. The reference image was registered to MNI space using FLIRT^68,69^ to estimate a subject-to-standard transformation, which was inverted using convert_xfm. The pretrained treatment transform weight image was transformed from MNI space into the subject- and visit-specific functional space using this inverse transformation and was used for all neurofeedback runs completed during that visit.

During each neurofeedback run, incoming volumes were motion corrected to the visit-specific reference image using MCFLIRT^68^. No spatial smoothing, temporal filtering, nuisance regression, or susceptibility-distortion correction was performed online. Each voxel was then standardized online using recursively updated estimates of the running mean and variance, following Welford’s algorithm. The standardized volume was multiplied voxelwise by the subject-space treatment transform mask, and the weighted voxel values were summed to yield a scalar treatment-space signal.

At the end of each scored scene, the transformed signal from all stations within that scene was concatenated and compared with the corresponding concatenated pre- and post-treatment templates. When a station volume was skipped, the corresponding participant and template timepoints were excluded from the correlations. Let 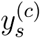 denote the concatenated treatment-space signal for participant *s* in scored scene *c*, and let 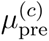 and 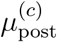 denote the corresponding scene-specific templates. Similarity to the post- and pre-treatment templates was quantified as

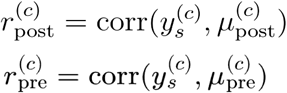

The displayed neurofeedback score was then computed as

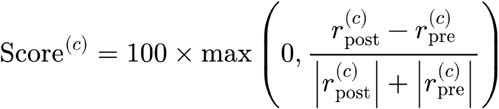

such that higher scores reflected greater similarity to the post-treatment template and lower similarity to the pre-treatment template. The denominator functioned to normalize scores across different scenes, which had variable overall correlation magnitudes. Negative contrasts were set to 0 to avoid discouraging feedback and ensure that any positive score indicated greater similarity to the post-treatment than the pre-treatment template. Scores were scaled to 0–100 and rounded to the nearest integer for display.

To preserve synchronization with movie playback, volumes were skipped when processing lag exceeded 4 seconds. Volumes acquired during the 22-second feedback interval were not processed, as these may have different mean and standard deviation from the movie volumes, disrupting the online standardization. This also allowed any lag between real-time processing and the movie stimulus to be reset after each feedback display. At each scored scene break, the analysis computer wrote the resulting score to a run-specific text file on the shared network drive. PsychoPy waited for this file during the 10-s “computing score” interval. On a small number of scored scenes, the participant instead saw a “could not compute score” display if more than half of the volumes falling within that scored scene were skipped. Sham participants received the corresponding score or missing-score display from their yoked real-neurofeedback participant. Participants were informed that missing scores resulted from technical issues and were unrelated to their brain activity.

### Post-task procedures

After exiting the scanner (within 45 minutes) participants completed a spontaneous speech task intended to probe the strategies they used and their experience with the novel neurofeedback design (see supplement for full instructions). As in our previous study,^34^ participants then completed questions related to comprehension of the movie narrative, recognition of specific objects in the movie, and overall quality of the movie audio and visuals. Participants also completed a scene-induced ratings survey, where, similar to our previous study,^34^ 3 second clips were selected from the movie, such that each of the 22 neurofeedback stations was represented, as well as any clips that contained drugs. Where possible, the clips were selected to overlap with clips used in our previous study and the peer reference sample. This yielded 33 clips in total; 13 clips contained depictions of opioid use or high; 12 clips were also rated by our peer reference sample; 6 clips met both criteria. Participants were instructed to watch each 3-second clip and provide subjective ratings of their experience when they had watched that clip in the scanner, including their craving for drugs, valence, arousal, and how closely related the scene is to their personal life experience. The order of the clips was randomized for subject and visit.

### MRI Data Acquisition

MRI scans were acquired using a Siemens 3.0 Tesla Skyra with a 32-channel head coil. Anatomical T1-weighted images were obtained using the following parameters: 3D Magnetization-Prepared Rapid Gradient-Echo (MPRAGE) sequence with 256 × 256 × 179 mm^3^ FOV, 0.8 mm isotropic resolution, TR/TE/TI = 2400/2.07/1000 ms, 8° flip angle with binomial (1, −1) fat saturation, 240 Hz/pixel bandwidth, 7.6 ms echo spacing, and in-plane acceleration [GRAPPA (generalized autocalibrating partially parallel acquisitions)] factor of 2, approximate acquisition time of 7 min. The blood-oxygen-level-dependent (BOLD) fMRI responses were measured as a function of time using T2*-weighted single-shot multiband accelerated (factor of 7) gradient-echo echoplanar image sequence [TE/TR = 35/1000 ms, 2.1 isotropic mm resolution, 70 axial slices without gaps for whole-brain coverage (147 mm), FOV 206 × 181 mm, matrix size 96 × 84, 60° flip angle (approximately Ernst angle), blipped CAIPIRINHA (Controlled Aliasing in Parallel Imaging Results in Higher Acceleration) phase-encoding shift = FOV/3, 1860 kHz/pixel bandwidth with ramp sampling, echo spacing 0.68 ms, and echo train length 84 ms].

### Anatomical preprocessing for offline analyses

Anatomical preprocessing was done with fMRIprep version 20.2.1. For each subject, a T1-weighted (T1w) image was corrected for intensity non-uniformity with N4BiasFieldCorrection, distributed with ANTs 2.3.3 and used as T1w-reference throughout the workflow. The T1w-reference was then skull-stripped with a Nipype implementation of the antsBrainExtraction.sh workflow (from ANTs), using OASIS30ANTs as the target template. Brain tissue segmentation of CSF, white matter and grey matter was performed on the brain-extracted T1w using FAST (FSL 5.0.9, RRID:SCR_00282339). Volume-based spatial normalization to a standard space (MNI152NLin2009cAsym) was performed through non-linear registration with antsRegistration (ANTs 2.3.3), using brain-extracted versions of both T1w reference and the T1w template. The ICBM 152 Nonlinear Asymmetrical template version 2009c (RRID:SCR_008796; TemplateFlow ID: MNI152NLin2009cAsym) was used for spatial normalization.

### Functional preprocessing for offline analyses

Initial functional preprocessing was done with fMRIprep version 20.2.1. For all BOLD neurofeedback runs (across all subjects), the following preprocessing was performed. First, a reference volume and its skull-stripped version were generated by aligning and averaging a single-band reference. A B0 non-uniformity map (or fieldmap) was estimated based on two EPI references with opposing phase-encoding directions, with AFNI’s 3dQwarp. Based on the estimated susceptibility distortion, a corrected EPI reference was calculated for a more accurate co-registration with the anatomical reference. The BOLD reference was then co-registered to the T1w reference using FLIRT^68,69^ (FSL 5.0.9) with the boundary-based registration cost-function. Co-registration was configured with nine degrees of freedom to account for distortions remaining in the BOLD reference. Head-motion parameters with respect to the BOLD reference (transformation matrices and six corresponding rotation and translation parameters) were estimated before any spatiotemporal filtering using MCFLIRT^68^ (FSL 5.0.9). The BOLD time-series were resampled onto their original, native space by applying a single, composite transform to correct for head-motion and susceptibility distortions. The BOLD time-series were resampled into the MNI152NLin2009cAsym standard space and used for further custom preprocessing.

### Custom preprocessing and parcellation for offline analyses

Spatially normalized, preprocessed BOLD data in the MNI152NLin2009cAsym standard space were further processed with the following steps. A grey matter mask was generated by averaging tissue probability maps of all subjects, then binarizing the resulting map by thresholding at 20% probability grey matter. The grey matter mask and Gaussian smoothing (6 mm) were then applied. The following confounds were regressed out of each BOLD signal: six translation and rotation parameters (x, y, z for each), their square, their derivative and their squared derivative, as well as the CSF component output by fMRIPrep. Regression of confounds, high-pass filtering (period of 140 s), and linear detrending were applied in a single step using the signal.clean function from nilearn Python package. Cortical regions of interest (ROI) were then delineated by applying the Schaefer functional parcellation with 1000 regions and 7 networks. Subcortical ROIs were selected with the Harvard-Oxford Subcortical Atlas with 20 regions.

To isolate movie-driven BOLD activity, the parcellated time series were trimmed to remove the first 34 volumes (20 s onset delay, 10 s to eliminate large stimulus onset responses, and 4 s to account for hemodynamic response lag) as well as volumes acquired during each 22-second inter-scene pause. Within each scan, the remaining segments were concatenated along the time axis to yield a continuous time series spanning the full movie. To isolate station-driven BOLD activity, the parcellated time series were trimmed to retain only volumes corresponding to neurofeedback stations (see Methods section *Feedback stations*) and concatenated along the time axis within each scan to yield a continuous time series spanning the stations alone. For both the whole-movie and station-only signals, each scan’s time series was z-scored along the time axis after trimming.

### Neurofeedback performance

To assess neurofeedback learning across runs, we fit a linear mixed-effects model predicting feedback performance from run, group (real, sham), and their interaction, with by-subject random intercepts and slopes and by-pair random intercepts (lme4/lmerTest in R; REML estimation with Satterthwaite degrees of freedom). Feedback performance was averaged across the 8 scene-level feedback scores within a run, and run was modeled as a continuous linear predictor centered at the first run, so the group main effect estimates the baseline (run-1) difference in scores. This model was fit to the first 4 runs and also assessed for the full 8 runs. We corrected across both run windows using the FDR approach detailed in the Results section *Neurofeedback learning and associated improvements in craving and affect*. Given our directional hypotheses that real feedback would yield a positive learning slope and steeper learning than sham, we used one-tailed tests. Fixed effects are reported as standardized regression coefficients (β), and effect sizes are reported as Cohen’s d.

### Craving and affective ratings

To assess craving changes over the course of training, we used responses from the scene-induced ratings survey (see Methods section *Post-task procedures*). To isolate craving-specific responses, we regressed out scene-induced valence and arousal ratings from the craving ratings within each subject, across sessions, and restricted the analyses to heroin-related clips. We then computed a weighted composite craving score as a weighted average of clip ratings, with weights derived to maximize the treatment effect in the peer reference sample (analogous to the treatment transform derived for the brain data; e.g., clips associated with the largest craving reductions in the peer reference sample received higher weights). This composite was used for all subsequent craving analyses. Scores were aligned to runs 1, 4, and 8, and within-subject deltas were computed between runs (4-1, 8-1). Four subjects (2 real, 2 sham) who completed runs 1–4 within a single session (precluding the computation of deltas) were excluded yielding a final sample of 16 subjects (8 real, 8 sham) for run 1-4 analyses. All of the below tests are included in the FDR correction approach detailed in the Results section *Testing PG-MovieNF efficacy*.

We tested whether real neurofeedback training reduced craving using a linear mixed-effects model with group (real, sham), run, and their interaction as fixed effects, and subject as a random intercept. Because the above exclusions left four subjects without yoked partners, pair was not included as a random effect in the primary model. To examine whether neurofeedback learning was associated with scene-induced craving reduction, we ran Spearman correlations between within-subject changes in feedback scores and changes in craving (run 4-1 and run 8-1 deltas).

Affective state was assessed using the Positive and Negative Affect Schedule (PANAS)^39^ at baseline (prior to run 1) and the final session (after run 8). A combined affective score was computed as the difference between positive and negative affect subscales (positive − negative). To test whether real PG-MovieNF altered affective state, we fit a linear mixed-effects model with group (real, sham), run (1, 8), and their interaction as fixed effects, and subject and yoked pair as crossed random intercepts. Effect sizes are reported as Cohen’s d. To examine the relationship between neurofeedback learning and affective change, we conducted Spearman correlations between feedback score deltas (run 8 − run 1) and positive > negative affect deltas (run 8 − run 1). Given our hypothesis that real PG-MovieNF would lead to a reduction in craving and improvement in affective ratings, we reported one-sided p-values.

### Shared orbitofrontal responses and target alignment

To test whether shared neural responses in orbitofrontal cortex (OFC) were temporally coupled with neurofeedback-related alignment dynamics, we developed a subject-level correlation approach linking each group’s shared responses to individual neurofeedback signals. These analyses were performed on the fully preprocessed BOLD data following the steps described above (see Methods sections *Anatomical preprocessing, Functional preprocessing,* and *Custom preprocessing and parcellation*). Because this pipeline included additional processing beyond the minimal steps applied to the real-time signal, we first verified that the neurofeedback learning effect remained intact. Recomputing the feedback scores offline not only replicated the real group’s real-time learning effect in the preprocessed data (*p* = .019), but also revealed a significant between-group difference in learning across runs 1–4 (*p* = .008), which was not observed in the real-time data (Supplementary Fig. S3; see Supplemental section *Offline Neurofeedback Scores*).

We first applied the Shared Response Model (SRM)^70^ using BrainIAK (v0.12) to estimate shared neural dynamics in the OFC. The OFC was defined as the set of Schaefer 1000 parcels falling within an anatomical OFC mask, excluding any parcels overlapping with the neurofeedback target mask. Parcellated BOLD time series were extracted for each parcel, subject, and run, then z-scored across time. For each subject, the remaining N − 1 subjects within the same group (real or sham) were used to fit a single-component SRM. Each subject’s input was a matrix of dimensionality p × t, where p is the number of OFC parcels and t is the number of TRs, averaged across runs 1–8 to produce one matrix per subject. The model learns subject-specific weight matrices that project multi-parcel OFC data into a common low-dimensional space, yielding a one-dimensional shared time course capturing the dominant pattern of temporally coordinated activity across OFC parcels that is common to each group. This leave-one-out shared response indexed the group-level OFC dynamics for each held-out subject. The SRM was fit with 20 iterations and a fixed random seed.

The alignment dynamics were computed as the element-wise product of each subject’s 1D transformed neurofeedback signal and the delta target signal at each TR. Positive values indicate moments when the subject’s brain activity is moving in the target direction; negative values indicate movement away from the target pattern. Alignment dynamics were computed separately for each target session (post-treatment, pre-treatment), and the final alignment signal for each subject’s run was defined as the difference (Δ = post − pre).

For each subject, we computed a Pearson correlation between the leave-one-out shared OFC response and a delta (post-treatment - pre-treatment) alignment signal. The shared response was paired with each run’s delta alignment signal across all eight neurofeedback runs, concatenated across runs, and correlated to yield a single r value per subject reflecting the degree to which fluctuations in the group’s shared OFC response tracked moment-to-moment changes in that subject’s neurofeedback alignment. This analysis was conducted separately for whole-movie and station-only time courses. For station-only analyses, the target signal was sliced at the same movie-relative time windows used to define the stations and concatenated to match the station-only transformed signal.

To test whether shared response–alignment coupling differed between groups, we conducted unpaired t-tests comparing real and sham groups. To test whether coupling was significantly present within each group independently, we conducted one-sample t-tests evaluating whether subject-level correlation coefficients differed from zero within the real and sham groups separately. To test whether the degree of shared response–alignment coupling tracked with a reduction in craving, we conducted a Spearman correlation between the shared response ∼ alignment correlation coefficients and the craving deltas (runs 4-1 and 8-1). Given our strong directional hypothesis of positive OFC coupling, we employed one-sided tests. These tests are included in the FDR approach detailed in the Results section *Testing PG-MovieNF efficacy*.

To examine the temporal relationship between shared OFC responses and alignment dynamics, we fit bivariate vector autoregressive models for each subject at the whole-movie level and tested Granger causality in both directions via F-tests. Stationarity was confirmed for all subjects using augmented Dickey–Fuller tests (all ps < 0.001). Lag order was selected per subject via minimizing the Akaike information criterion (AIC). Because AIC-selected lag orders differed between groups (t(18) = 2.15, p = 0.046; real mean = 19.7, sham mean = 17.0), we also repeated the analysis with a fixed lag equal to the median across all subjects (lag = 20), which did not change results (real: *t*(9) = 3.15, *p* = 0.012; between-group: *t*(18) = 2.14, *p* = 0.046). Group-level inference was conducted using one-sample t-tests on subject-level F-statistics and between-group comparisons using unpaired t-tests.

## Data availability

The data supporting the findings of this study are available from the corresponding authors upon reasonable request.

## Code availability

The real-time fMRI analysis pipeline and PsychoPy implementation of the PG-MovieNF task are publicly available on GitHub: https://github.com/brainiak/rtcloud-projects/tree/main/movienf.

## Author contributions

R.Z.G. conceived the study. G.K., N.E.M., A.C.H., U.H., K.A.N. and R.Z.G. designed the study. G.K. developed the software. G.K., N.E.M., M.B., S.S. and K.R.D. collected the data. G.K. and N.E.M. performed the analyses. N.E.M. and G.K. prepared the figures. G.K., N.E.M. and R.Z.G. wrote the original draft. G.K., N.E.M., K.A.N., R.Z.G., U.H., A.C.H. and N.A.-K. reviewed and edited the manuscript. R.Z.G., K.A.N., N.A.-K. and U.H. supervised the research. R.Z.G. secured funding.

