## Supplementary Information for "Peer-guided movie neurofeedback to facilitate recovery in opioid use disorder"

### Peer Reference Sample

Demographic, clinical, and substance use measures for the peer reference sample (N = 37) are summarized in Table S1 (see Supplemental section *Eligibility Criteria, Diagnostic Interviews, and Baseline Assessments* for more details on these measures). As expected, heroin abstinence increased significantly from pre- to post-treatment ($p$ < .001). No other measures differed significantly between sessions ($ps \geq .051$).

| | Pre-Treatment | Post-Treatment | sig. test |
|---|---|---|---|
| Age | 43.08 (10.82) | | |
| Sex (Male/Female) | 30/7 | | |
| Race (White/Black/Other) | 24/5/8 | | |
| Ethnicity (Hispanic/Non-Hispanic) | 13/24 | | |
| Education (years) | 12.00 (2.37) | | |
| Verbal IQ | 98.11 (10.63) | | |
| Nonverbal IQ | 10.54 (3.02) | | |
| Handedness (Right/Left) | 29/8 | | |
| Beck Depression Inventory (BDI) | 11.54 (8.58) | 9.25 (8.87) | 0.051 |
| Beck Anxiety Inventory (BAI) | 9.30 (8.74) | 8.68 (9.34) | 0.660 |
| Smoking Status (Current/Past/Never) | 35/2/0 | 33/4/0 | 0.674[a] |
| Fagerström Test for Nicotine Dependence (FTND) | 3.41 (1.85) | | |
| Short Michigan Alcoholism Screening Test (SMAST) | 3.14 (3.63) | | |
| Lifetime Heroin Use (years) | 14.25 (10.51) | | |
| Heroin Abstinence (days) | 191.14 (214.40) | 251.97 (243.57) | **<.001[b]** |
| Heroin Use Past Month (days) | 0.24 (0.72) | 0.70 (2.70) | 0.321[b] |
| Heroin Craving Questionnaire (HCQ) | 36.30 (13.36) | | |
| Subjective Opiate Withdrawal Scale (SOWS) | 3.95 (6.55) | | |
| Severity Of Dependence Scale (SDS) | 10.95 (4.16) | | |
| Methadone Dosage (mg) | 103.77 (63.57) | 107.75 (75.97) | 0.634[b] |

**Table S1. Peer reference sample characteristics by session.** Paired t-tests were conducted to assess session differences. Values in parentheses denote standard deviation. Bold typeface indicates significant between-session differences, correcting for familywise error (α=.05/6=.008). [a]Chi-square tests were used for unordered categorical and binary data. [b]Wilcoxon signed-rank tests were used when assumptions of normality were violated.

### Formal derivation for deltaCorrCA

The main Methods define the processed, parcellated BOLD data for subject $i$ and session $j \in \{\text{pre}, \text{post}\}$ as $X_{ij} \in \mathbb{R}^{R \times T}$, where $R$ is the number of regions of interest and $T$ is the number of repetition times. The treatment transform $W \in \mathbb{R}^{R}$ projects these multivariate responses into a one-dimensional treatment-space signal,

$y_{ij} = W^{\top} X_{ij}$

For the derivation below, all session-level treatment-space signals and templates are z-scored across time. For notational simplicity, the subscript $-s$ denotes the training subjects used to classify held-out subject $s$. In the ten-fold cross-validation analysis, the entire test fold containing subject $s$, including both sessions from every subject in that fold, was excluded from model fitting and template construction. For held-out subject $s$, we define the classifier score for a treatment-space signal $y$ as

$$g_s(y) = \mathrm{corr}(y, \mu_{\mathrm{post},-s}) - \mathrm{corr}(y, \mu_{\mathrm{pre},-s})$$

where $\mu_{\mathrm{pre},-s}$ and $\mu_{\mathrm{post},-s}$ are the pre- and post-treatment templates estimated from the training subjects. Positive values of $g_s(y)$ favor a post-treatment classification, whereas negative values favor a pre-treatment classification. Define the training-set template-delta signal as

$$\Delta\mu_{-s} = \mu_{post,-s} - \mu_{pre,-s}$$

and the held-out subject's projected treatment-delta signal as

$$y_{\Delta,s} = y_{s,post} - y_{s,pre}$$

The paired classifier margin is the extent to which the post-treatment session receives a more post-favoring score than the corresponding pre-treatment session:

$$m_s = g_s(y_{s,post}) - g_s(y_{s,pre})$$

Substituting the classifier score into the paired margin gives

$$\begin{aligned} m_s = {} & \mathrm{corr}(y_{s,\mathrm{post}}, \mu_{\mathrm{post},-s}) - \mathrm{corr}(y_{s,\mathrm{post}}, \mu_{\mathrm{pre},-s}) \\ & - \mathrm{corr}(y_{s,\mathrm{pre}}, \mu_{\mathrm{post},-s}) + \mathrm{corr}(y_{s,\mathrm{pre}}, \mu_{\mathrm{pre},-s}) \end{aligned}$$

Because the session-level signals and templates are z-scored across time, Pearson correlation between two length-$T$ time series $a$ and $b$ can be written as

$$\mathrm{corr}(a, b) = \frac{1}{T-1} a b^{\top}$$

The paired margin can therefore be written in terms of dot products as

$$\begin{aligned} m_s = \frac{1}{T-1} \Big[ & y_{s,\mathrm{post}} \mu_{\mathrm{post},-s}^{\top} - y_{s,\mathrm{post}} \mu_{\mathrm{pre},-s}^{\top} \\ & - y_{s,\mathrm{pre}} \mu_{\mathrm{post},-s}^{\top} + y_{s,\mathrm{pre}} \mu_{\mathrm{pre},-s}^{\top} \Big] \end{aligned}$$

Regrouping terms gives

$$m_s = \frac{1}{T-1} \left(y_{s,post} - y_{s,pre}\right)\left(\mu_{post,-s} - \mu_{pre,-s}\right)^{\top}$$

Using the definitions above,

$$m_s = \frac{1}{T-1} y_{\Delta,s} \Delta\mu_{-s}^{\top}$$

Both $y_{\Delta,s}$ and $\Delta\mu_{-s}$ are centered across time because they are differences between centered time series. Their inner product can therefore be expressed in terms of their Pearson correlation:

$$m_s = \kappa_s \,\mathrm{corr}(y_{\Delta,s}, \Delta\mu_{-s})$$

where

$$\kappa_s = \frac{\|y_{\Delta,s}\|_2 \|\Delta\mu_{-s}\|_2}{T-1} > 0$$

The above correlation is equivalent to the leave-one-subject-out treatment-delta ISC:

$$\mathrm{ISC}_s(W) = \mathrm{corr}(y_{\Delta,s}, \Delta\mu_{-s})$$

Thus, the paired classifier margin can be written directly in terms of treatment-delta ISC:

$$m_s(W) = \kappa_s(W)\,\mathrm{ISC}_s(W)$$

Because $\kappa_s(W)$ is strictly positive, the classifier margin and treatment-delta ISC have the same sign. DeltaCorrCA identifies a common spatial projection for which these post-minus-pre movie dynamics are reliably shared across subjects, i.e., maximizing the treatment delta ISC. This is the same structure rewarded by the classifier margin: the margin becomes positive when a held-out participant's projected treatment delta follows the temporal direction expressed by the peer-derived template delta. DeltaCorrCA therefore learns a treatment-space dimension that emphasizes reproducible change across peers, while cross-validated template matching tests whether that shared treatment direction generalizes to unseen participants. Together, these steps convert longitudinal treatment-related change in the peer reference cohort into a neural target that can be applied during neurofeedback.

### Candidate Models Evaluated During Target Identification

To identify the ROI set and parameters used for the PG-MovieNF treatment transform, we performed a grid search over candidate ROI sets. These candidate sets were defined a priori from anatomical and functional atlases (e.g., Brodmann areas, Schaefer parcellation labels, Harvard-Oxford atlas labels and canonical networks in the addiction literature) together with combinations of these definitions. For each ROI set, we also tested 4 values for the CorrCA regularization parameter (0, 0.1, 0.2, 0.5). We additionally tested models with and without overall mean subtraction, where the mean over all subjects and sessions within each ROI is first subtracted out to remove any signal that is common to both pre and post treatment. For each combination, the deltaCorrCA model was fit to the within-subject post-minus-pre treatment difference signals and evaluated by 10-fold cross-validated classification accuracy of pre- versus post-treatment labels, which was used to rank tested models (see Table S2). We note that mean subtraction improved performance for all ROI sets, so we do not report that parameter in Table S2. The top-performing model used bilateral dlPFC ROIs and part of the bilateral dmPFC (defined by Brodmann areas 9 and 46) with a regularization of 0.1 and mean subtraction, achieving a mean held-out accuracy of 0.775, supporting a focal dlPFC/dmPFC target for the treatment transform. This model was carried forward and refit on all 37 peer reference iOUD to derive the final transform (W) and pre/post templates for PG-MovieNF training.

| Rank | ROI set | ROI definition | Mean held-out accuracy | CorrCA regularization |
|---|---|---|---|---|
| 1 | dl/dmPFC | Brodmann areas 9 and 46 | 0.775 | 0.1 |
| 2 | dl/dmPFC + vlPFC | Union of dl/dmPFC and vlPFC ROI sets | 0.763 | 0.1 |
| 3 | vlPFC | Brodmann areas 44, 45, and 47 | 0.750 | 0.1 |
| 4 | broad medial/lateral PFC + subcortical | Union of dmPFC, vmPFC, dlPFC, vlPFC, and Harvard-Oxford subcortical ROIs: striatum, hippocampus, and amygdala | 0.738 | 0.2 |
| 5 | broad medial/lateral PFC | Union of dmPFC, vmPFC, dlPFC, and vlPFC ROI sets | 0.738 | 0.2 |
| 6 | salience/limbic | Union of Schaefer salience/ventral attention and limbic network ROIs | 0.725 | 0.1 |
| 7 | non-sensory cortex | All cortical ROIs excluding sensory-network ROIs | 0.725 | 0.0 |
| 8 | control/limbic/subcortical | Union of Schaefer control and limbic network ROIs, plus Harvard-Oxford striatum, hippocampus, and amygdala | 0.700 | 0.0 |
| 9 | dmPFC | Brodmann areas 9, 10, and 32 | 0.700 | 0.2 |

| | | | | |
|---|---|---|---|---|
| 10 | salience/ventral attention network | Schaefer salience/ventral attention network ROIs | 0.700 | 0.0 |
| 11 | salience/control /limbic/subcortical | Union of Schaefer salience/ventral attention, control, and limbic network ROIs, plus Harvard-Oxford striatum, hippocampus, and amygdala | 0.688 | 0.5 |
| 12 | ventral striatum | Harvard-Oxford accumbens ROIs | 0.688 | 0.1 |
| 13 | dorsal attention network | Schaefer dorsal attention network ROIs | 0.675 | 0.2 |
| 14 | dmPFC/vmPFC | Union of dmPFC and vmPFC ROI sets | 0.675 | 0.5 |
| 15 | iRISA network union | Union of OFC, vmPFC, insula, ACC, PCC, precuneus, dlPFC, vlPFC, dmPFC, and Harvard-Oxford subcortical ROIs: striatum, hippocampus, and amygdala | 0.675 | 0.0 |
| 16 | salience/control /limbic | Union of Schaefer salience/ventral attention, control, and limbic network ROIs | 0.663 | 0.5 |
| 17 | default/limbic | Union of Schaefer default mode and limbic network ROIs | 0.625 | 0.2 |
| 18 | all cortical ROIs | All Schaefer cortical ROIs | 0.625 | 0.1 |
| 19 | insula | Schaefer insula parcels and Harvard-Oxford insula ROIs | 0.613 | 0.0 |
| 20 | default/control | Union of Schaefer default mode and control network ROIs | 0.613 | 0.5 |
| 21 | vmPFC | Brodmann areas 10, 11, 12, 25, and 32 | 0.600 | 0.5 |
| 22 | striatum | Harvard-Oxford caudate, putamen, pallidum, and accumbens ROIs | 0.575 | 0.5 |
| 23 | frontoparietal control network | Schaefer control network ROIs | 0.563 | 0.5 |
| 24 | OFC/vmPFC | Schaefer OFC parcels, Brodmann areas 10, 11, 12, 25, 32, and 47, and Harvard-Oxford frontal orbital cortex | 0.563 | 0.0 |
| 25 | default mode network | Schaefer default mode network ROIs | 0.563 | 0.2 |
| 26 | OFC | Schaefer OFC parcels and Harvard-Oxford frontal orbital cortex | 0.538 | 0.1 |
| 27 | cortical limbic network | Schaefer limbic network ROIs | 0.525 | 0.1 |

**Table S2. Models tested for target identification, ranked by cross-validated classification accuracy.** Each row is a candidate model defined by an ROI set and its definition (Brodmann areas, Schaefer atlas labels, Harvard-Oxford labels, or unions thereof). Mean held-out accuracy is the mean classification accuracy across folds for classifying pre- versus post-treatment sessions on held-out subjects, in a 10-fold cross-validation setup. CorrCA regularization is the shrinkage parameter applied when estimating the correlated component. The top model was selected for derivation of the PG-MovieNF target signal and treatment transform.

### Late-run task disruption motivates testing runs 1–4

Because this was the first implementation of peer-guided movie neurofeedback in this population, we conservatively chose a large number of runs to reduce the risk that low performance would simply reflect insufficient opportunity to practice the task. At the same time, extending training across repeated exposures to the same movie introduced the opposite possibility: later runs could become increasingly influenced by declining engagement and real-time processing delays, making them less interpretable as a pure measure of neurofeedback learning. We therefore asked whether the full eight-run trajectory was better characterized as an early learning phase followed by a later phase increasingly affected by task disruption, and whether this distinction justified focusing analyses on an early learning phase.

To avoid choosing an arbitrary cutoff in advance, we first modeled the real-group feedback trajectory continuously across all eight runs. A random-intercept cubic mixed-effects model fit the data better than both a linear model (likelihood-ratio test: LR = 7.39, $p = 0.025$) and a quadratic model (LR = 6.08, $p = 0.014$). The fitted cubic curve identified an early local maximum at approximately run 2.94 and a later local minimum at approximately run 6.66, consistent with an initial learning phase followed by a late drop in scores (Figure S1A). We then fit a piecewise linear mixed model with a prespecified knot at run 4. This model yielded a positive early slope ($\beta = 2.49$, one-sided $p = 0.068$) and a negative late slope ($\beta = -2.31$, one-sided $p = 0.030$), with a negative post-run-4 slope shift ($\beta = -4.80$, one-sided $p = 0.032$). Thus, two distinct model classes converged on the same qualitative pattern: scores improved during the early runs and declined after run 4. The early slope in the piecewise model did not itself reach significance; the significant learning effect across runs 1–4 comes from the mixed-effects model reported in the main text.

We next asked whether the late-run decline could plausibly reflect factors relating to task disruption rather than the intended learning process. Task engagement ratings were treated as an a priori disruption variable, whereas the number of skipped TRs were identified post hoc as a technical factor. Network-related processing delays occasionally caused the real-time analysis pipeline to fall behind movie playback, requiring incoming TRs to be skipped to preserve synchronization between the analysis and the movie. Skipped TRs occurring during stations can affect the accuracy of the delivered feedback and excessive skipped TRs prevented a score from being displayed at all ("could not compute score" displayed, see Methods, *Neurofeedback Task Design*), both of which we expected to be disruptive to learning. Although there was no a priori reason to expect elevated skipped-TR rates to emerge specifically after run 4, quality control revealed a peak in skipped TRs during runs 4-6 (Figure S1B), which coincided with the observed drop in scores after run 4. The average number of skipped TRs was significantly higher for runs 4-6 compared to all other runs ($\beta = 0.055$, 95% CI [-0.007, 0.116], one-sided $p = 0.041$; Figure S1B). Confidence intervals in this section are two-sided, whereas tests are one-sided given directional hypotheses. Engagement declined linearly over runs ($\beta = -0.149$, 95% CI [-0.308, 0.011], one-sided $p = 0.034$) and was lower in runs 5–8 than in runs 1–4 ($\beta = -0.753$, 95% CI [-1.229, -0.278], one-sided $p = 0.0010$; Figure S1C). To summarize declining engagement and real-time processing delays in a single run-level measure, we defined a composite disruption index at run t as zscore(skipped_trs_fraction(t−1)) − zscore(engagement(t)), and then z-scored the resulting composite so that higher values indicated greater task disruption. This composite disruption index showed a clear upward shift beginning at run 5 ($\beta = 0.583$, 95% CI [0.133, 1.033], one-sided $p = 0.006$; Figure S1D).

We then tested whether the individual disruption indicators and the composite disruption index predicted reduced feedback scores. Because current run performance should depend partly on the previous run's score, we fit full-run autoregressive models in the real group with current feedback score as the outcome and previous-run feedback as a covariate. These repeated-measures models were fitted with subject-clustered Gaussian generalized estimating equation (GEE) models using an exchangeable working correlation structure[1]. We used GEE rather than comparable mixed-effects models because mixed-effects models were sometimes singular. Because these analyses tested directional hypotheses, one-sided p-values are reported for the inferential tests below, whereas standard two-sided 95% confidence intervals are retained to describe effect size and uncertainty.

Across the full run series, higher skipped-TR fraction on the previous run predicted lower current feedback score when entered alone in an autoregressive model ($\beta = -2.25$, 95% CI [-4.76, 0.26], one-sided $p = 0.0397$). Current post-run engagement also predicted higher current feedback when entered alone ($\beta = 3.22$, 95% CI [0.20, 6.25], one-sided $p = 0.018$). When previous run skipped TR fraction and current engagement were entered together, the skipped-TR effect remained significant ($\beta = -2.50$, 95% CI [-4.81, -0.19], one-sided $p = 0.017$), whereas the current-engagement effect remained positive but was significant only at trend level ($\beta = 2.62$, 95% CI [-0.65, 5.89], one-sided $p = 0.058$; Figure S1E). The composite disruption index was an even stronger predictor of current feedback across all runs ($\beta = -3.78$, 95% CI [-5.59, -1.98], one-sided $p = 0.00002$; Figure S1E). These autoregressive results indicate that scores across the task were sensitive not only to prior performance, but also to previous-run real-time processing delays and current-run engagement. In particular, the lagged skipped-TR effect supports the interpretation that real-time processing delays interfered with learning and reduced scores on the following run.

Finally, we tested whether the same disruption variables explained the late drop below the end of the early learning period. For each participant, late-run change was calculated by subtracting their run-4 performance from their performance in each of runs 5–8, such that more negative values indicated a larger decline relative to run 4. Run number was included as a covariate to account for systematic change across the late-run period. Current engagement robustly predicted less late drop ($\beta = 5.26$, 95% CI [2.04, 8.48], one-sided $p = 0.0007$; Figure S1F), whereas previous-run skipped-TR fraction alone did not significantly predict late-drop magnitude in the hypothesized direction. The composite disruption index, however, significantly predicted greater drop below run-4 performance ($\beta = -4.23$, 95% CI [-7.91, -0.56], one-sided $p = 0.012$; Figure S1F).

Taken together, these analyses suggest that the decline beginning at run 5 is not best interpreted simply as an absence of further learning. Rather, later-run scores were increasingly affected by task disruption. Engagement, which we expected a priori to become more limiting over repeated training, declined over runs and predicted both current score and the extent of late drop. Independently, the skipped-TR fraction peaked around runs 4–6 and predicted lower subsequent scores in autoregressive models, consistent with the idea that real-time processing delays interfered with learning and degraded performance on the next run. The composite disruption index provided the strongest and most parsimonious summary of these effects, predicting both lower current scores across the run series and greater late drop below run 4. These results indicate that runs 5–8 were increasingly influenced by real-time processing delays and declining engagement and were therefore less clean measures of neurofeedback learning than runs 1–4. At the same time, there may be effects of additional neurofeedback training despite the disruption, and several downstream outcomes, including craving and affective ratings, were collected after the full 8-run protocol. We therefore test both windows of runs 1-4 and runs 1-8 for the primary analyses where data is available.

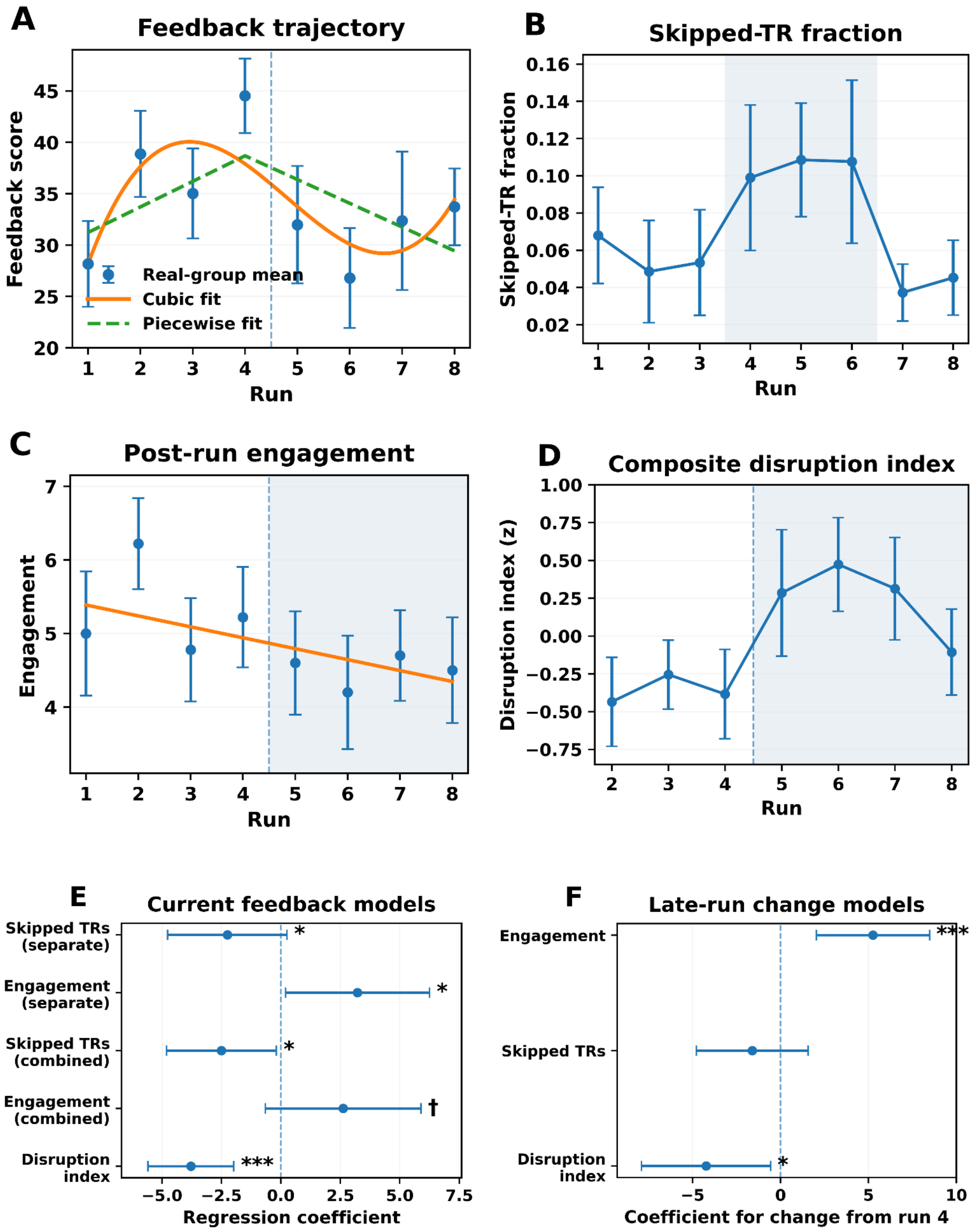


**Figure S1. Late-run task disruption and rationale for emphasizing runs 1–4.**
**(A)** Real-group feedback trajectory across eight runs. Points show run means ± s.e.m.; the solid and dashed curves show the cubic mixed-effects and piecewise linear fits, respectively. The cubic model fit better than the linear (LR = 7.39, $p$ = .025) and quadratic models (LR = 6.08, $p$ = .014), with an estimated peak at run 2.94 and trough at run 6.66. In the piecewise model, the early slope was positive at trend level (β = 2.49, one-sided $p$ =

.068), the late slope was negative (β = −2.31, one-sided $p$ = .030), and the post-run-4 slope shift was negative (β = −4.80, one-sided $p$ = .032). The vertical dashed line marks the transition after run 4.
**(B)** Skipped-TR fraction across runs. The shaded region marks runs 4–6, during which skipped-TR fraction was higher than during the remaining runs (β = .055, 95% CI [−.007, .116], one-sided $p$ = .041).
**(C)** Post-run engagement declined across runs (β = −.149, 95% CI [−.308, .011], one-sided $p$ = .034) and was lower during runs 5–8 than runs 1–4 (β = −.753, 95% CI [−1.229, −.278], one-sided $p$ = .0010). The shaded region marks runs 5–8.
**(D)** Disruption index across runs. The index was defined as $z$(previous-run skipped-TR fraction) − $z$(current engagement) and then z-scored, such that higher values indicate greater previous-run processing delays together with lower current engagement. Note that the disruption index is not defined for run 1 as there is no previous run available for the skipped TR fraction. The disruption index increased beginning at run 5 (β = .583, 95% CI [.133, 1.033], one-sided $p$ = .006).
**(E)** Coefficients and two-sided 95% confidence intervals from autoregressive GEE models predicting current feedback while adjusting for previous-run feedback. In separate models, previous-run skipped TRs predicted lower current feedback (β = −2.25, 95% CI [−4.76, .26], one-sided $p$ = .0397), whereas current engagement predicted higher feedback (β = 3.22, 95% CI [.20, 6.25], one-sided $p$ = .018). In the combined model, the skipped-TR effect remained significant (β = −2.50, 95% CI [−4.81, −.19], one-sided $p$ = .017), whereas engagement remained positive at trend level (β = 2.62, 95% CI [−.65, 5.89], one-sided $p$ = .058). The disruption index was a strong predictor of lower current feedback (β = −3.78, 95% CI [−5.59, −1.98], one-sided $p$ = .00002).
**(F)** Coefficients and two-sided 95% confidence intervals from GEE models predicting feedback during runs 5–8 relative to each participant's run-4 score while adjusting for centered late-run time. Current engagement predicted less late-run decline (β = 5.26, 95% CI [2.04, 8.48], one-sided $p$ = .0007), skipped TRs did not significantly predict late-run decline (β = −1.61, 95% CI [−4.79, 1.57], one-sided $p$ = .160), and the disruption index predicted greater decline (β = −4.23, 95% CI [−7.91, −.56], one-sided $p$ = .012). Symbols are shown only in E–F and indicate †$p$ < .10, *$p$ < .05, **$p$ < .01, and ***$p$ ≤ .001. Error bars in A–D indicate s.e.m.; horizontal lines in E–F indicate two-sided 95% confidence intervals. Unless otherwise stated, inferential $p$-values are one-sided for directional hypotheses.

### Effects of deviations from the intended run structure

Although the intended training schedule was two runs per session, 10 of 20 participants followed that schedule exactly through run 4, and 5 of 20 followed it exactly through run 8. To test whether session placement affected learning beyond cumulative practice, we fit separate linear mixed-effects models across runs 1–4 and 1–8. Extending the main-text model, fixed effects included group, cumulative run, actual session, group × cumulative run, and group × actual session; random effects included correlated participant-specific intercepts and run slopes and a yoked-pair intercept. The run coefficient therefore tested improvement per additional run while holding session constant, whereas the session coefficient tested whether the same cumulative run differed when completed one session later. Across runs 1–4, the session-adjusted run effect was significant in the real group (β = .38, t(28.6) = 1.81, one-sided p = .040), whereas session placement was not (β = .02, t(31.9) = 0.13, two-sided p = .898). Across runs 1–8, neither cumulative run (β = −.001, t(72.7) = −0.002, one-sided p = .501) nor session placement (β = −.07, t(64.9) = −0.27, two-sided p = .791) was significant. Neither effect differed by group in either window (group × run, one-sided ps ≥ .197; group × session, two-sided ps ≥ .459). Thus, the early improvement reported in Figure 3A remained after accounting for session placement, whereas reallocating runs across sessions did not detectably influence performance. Directional run-learning effects were tested one-sided, consistent with the main analyses; session-placement effects were tested two-sided.

### Controlling for baseline craving in the craving reduction models

To test whether the group difference in craving reduction was attributable to baseline differences in craving prior to neurofeedback training, we assessed group differences in pre-task craving measures and their contributions to the group × run interaction effects. Two baseline craving measures were assessed: 1) a Likert rating of imagined likelihood of heroin use if placed in a previous drug-use environment, a measure of cue-induced craving, collected in the scanner immediately before the first run (the more state-dependent measure); and 2) the Heroin Craving Questionnaire (HCQ), a trait-level measure completed outside the scanner prior to neurofeedback training. These two measures did not differ between groups at baseline (imagined craving: $t(17.9) = -1.34$, $p = .197$; HCQ: $t(17.7) = -0.11$, $p = .913$). Moreover, adjusting for baseline craving did not change the between-group craving reduction effects. Controlling for imagined craving, the group × run interaction remained significant for both runs 1-4 ($\beta = 0.73$, $t(14) = 2.78$, $p = .007$, $d = 1.96$) and runs 1–8 ($\beta = 0.66$, $t(55) = 3.02$, $p = .002$, $d = 1.91$). The same effect held when controlling for HCQ (runs 1–4: $\beta = 0.73$, $t(14) = 2.78$, $p = .007$, $d = 1.96$; runs 1–8: $\beta = 0.66$, $t(55) = 3.03$, $p = .002$, $d = 1.91$). These estimates are identical to the unadjusted model (runs 1–4: $\beta = 0.73$, $t(14) = 2.78$, $p = .007$; runs 1–8: $\beta = 0.66$, $t(56) = 3.05$, $p = .002$), indicating that the group difference in craving reduction is not confounded by baseline differences in craving.

### Synchronization in OFC dynamics and alignment dynamics

In addition to assessing the degree of coupling between the OFC and alignment dynamics, we also assessed the ISC within each of these signals alone during feedback stations across runs. Shared OFC responses were significantly synchronized in both groups (real: $M = 0.262$, $t(9) = 8.14$, $p < .001$; sham: $M = 0.163$, $t(9) = 6.40$, $p < .001$), and were more synchronized in the real than the sham group ($t(18) = 2.41$, $p = .028$). The alignment signals were also significantly synchronized across subjects within each group (real: $M = 0.065$, $t(9) = 4.17$, $p = .002$; sham: $M = 0.075$, $t(9) = 6.48$, $p < .001$), with no group difference (Welch's $t(18) = -0.52$, $p = .61$).

### Real PG-MovieNF increases prefrontal inter-subject synchronization

To assess whether PG-MovieNF training induced convergent neural dynamics across participants, we examined inter-subject correlation (ISC) across prefrontal cortex (PFC) ROIs over the course of training, proceeding from the 1D projected neurofeedback signal to a broader ROI-wise analysis across PFC parcels.

We first tested whether the 1D transformed neurofeedback signal itself became more synchronized across participants over training. ISCs were computed within each run and slopes across runs 1–4 were estimated with OLS regression per subject, then tested with one-sample t-tests and compared between groups with Welch's t-test. The sham group showed significantly negative slopes (sham: mean = −0.033, $t(9) = -2.59$, $p = .029$), while the real group showed a similar yet trending effect (real: mean = −0.027, $t(9) = -2.20$, $p = .056$); the groups did not differ from each other (Welch's $t = 0.32$, $p = .75$).

Given the absence of convergence at the level of the 1D signal, we conducted a broader ROI-wise analysis across 179 PFC parcels (Schaefer atlas). Leave-one-out ISCs were computed on preprocessed whole-movie BOLD data for each parcel and run, with each subject's time series correlated against the group mean of the remaining subjects and Fisher z-transformed prior to analysis. Per-subject ISC slopes across runs 1–4 were estimated with OLS regression and compared between groups using unpaired permutation tests (10,000 permutations), with p-values FDR-corrected ($q < .05$) across PFC parcels. Twenty-six parcels showed significant between-group differences in ISC slope, spanning frontoparietal control (13 ROIs), salience/ventral attention (10 ROIs), and limbic (3 ROIs) networks, with a left-hemisphere predominance (17 LH, 9 RH). In 25 of the 26 ROIs, the real group showed less negative slopes than the sham group (ds = 1.48–2.96), five of which overlapped with the dlPFC mask used to derive the neurofeedback signal. The largest effects were in left frontal opercular regions of the salience/ventral attention network (FrOper22_LH: Δ = +0.080, $p_{FDR}$ = .005, d = 2.96; FrOper13_LH: Δ = +0.035, $p_{FDR}$ = .005, d = 2.68). The one exception was a left orbitofrontal limbic parcel where the sham group showed a less negative slope (OFC2_LH: Δ = −0.030, $p_{FDR}$ = .011, d = −2.15).

Post-hoc within-group sign-flipped permutation tests identified the contributions of each group. In the real group, several left salience/ventral attention frontal opercular parcels showed significantly positive slopes, reflecting an increase in synchronization over training (ds = +1.03 to +1.29; e.g., FrOper18_LH: mean = +0.032, p = .003, d = 1.29), while two parcels showed significantly negative slopes (OFC2_LH: d = −1.43, p = .006; PFCl17_RH: d = −1.06, p = .010). In the sham group, 22 of the 26 ROIs exhibited significantly negative slopes, reflecting widespread PFC desynchronization over training (ds = −0.76 to −2.79, all ps < .030); the strongest effects were in bilateral lateral PFC within the frontoparietal control network (PFCl3_LH: mean = −0.066, p = .003, d = −2.79; PFCl17_RH: mean = −0.058, p < .001, d = −2.72). Together, the between-group differences were thus driven by a combination of focal synchronization increases in the real group and broad desynchronization in the sham group.

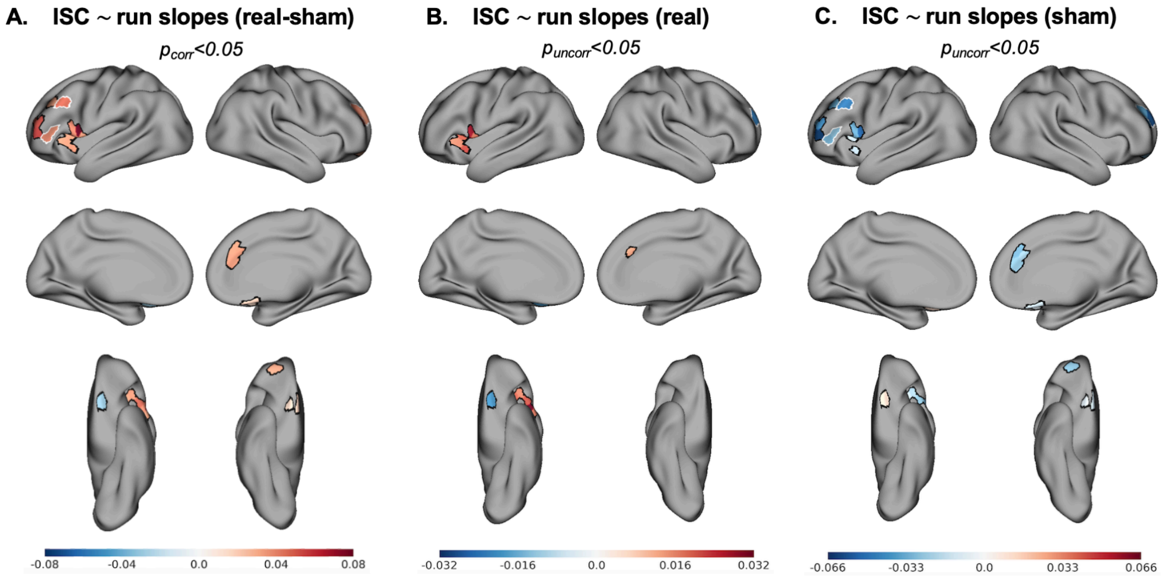


**Figure S2. Increased prefrontal synchronization in the real neurofeedback group. A.** Between-group differences in ISC run slopes (real − sham), thresholded at $p_{FDR}$<.05. Warm colors indicate more positive slopes in the real group; cool colors indicate more positive slopes in the sham group. Significant ROIs were concentrated in the left frontoparietal control and salience/ventral attention networks, with the largest effects in the left frontal opercular cortex. **B-C.** Post-hoc one-sample ISC run slopes within the real **B.** and sham **C.** groups, restricted to the 26 ROIs identified in **A.** and thresholded at $p_{uncorr}$<.05. In the real group, positive slopes (warm colors) emerged in salience/ventral attention frontal opercular regions, indicating increasing synchrony across runs. In the sham group, widespread negative slopes (cool colors) indicate progressive desynchronization across PFC. Color bars reflect mean run ~ ISC slope. ROIs outlined in white indicate those included in the dlPFC mask. Surface maps are displayed in lateral, medial, and ventral views.

### PG-MovieNF Task instructions

“In this task you will watch a movie about heroin addiction.
Your goal is to make your brain activity similar to a target brain activity pattern associated with positive treatment outcomes for addiction.

Your brain activity will be recorded and the movie will be paused at select times to present a feedback score (0-100) indicating how similar your brain was to the target during the previous scene
Use this feedback to learn a mental strategy that maximizes your scores and makes your brain activity as similar to the target as possible

Here we will offer some suggestions for mental strategies that could help maximize your feedback score.
However, you should use any strategy that you think may work for you.
For scenes involving drugs, you could try to reduce your emotional reaction. You could do so by focusing on taking slow deep breaths or reminding yourself that the scenario is not real and the drugs are not real. It is a movie and that these are actors.
Alternatively you could think about some negative consequences of using drugs or the positive consequences of remaining in treatment. Any other strategy that you think could decrease your emotional reaction to heroin-related content could be helpful.

Ultimately the strategy that you use is up to you.
The most important thing is to maximize your feedback scores.
Try to notice how your strategy, thoughts, or feelings affect your score and adjust accordingly.
If a strategy is consistently yielding low scores, try to change it.
If a strategy is consistently yielding high scores, stay with it.
After each training session we will ask you about the strategies you tried and how you think they impacted your scores, if at all."

### Post-NF Speech Task instructions

"Please describe your experience trying to maximize your neurofeedback score during today's session.
Describe any strategies, thoughts or feelings that you used to try to maximize your score during the movie.
Which strategies, if any, seemed to work and which did not? Why do you think they did or didn't work?
How did the feedback or your strategy affect your experience of the movie?
How do you feel in general about this experiment? Did it help you in any way?
You can tell us any positive or negative impressions you had."

### Eligibility Criteria, Diagnostic Interviews, and Baseline Assessments

All participants met the following inclusion criteria: 1) Ability to understand and give informed consent; and 2) 18-64 years of age. All iOUD further met the following inclusion criteria: 1) Diagnostic and Statistical Manual of Mental Disorders (DSM-5) diagnosis of OUD with heroin as the primary drug of choice; and 2) stabilized on medication for opioid use disorder (i.e., methadone or suboxone). Participants were excluded from the study if they met any of the following criteria: 1) DSM-5 diagnosis for schizophrenia or developmental disorder (e.g., autism); 2) head trauma with loss of consciousness (>30 min); 3) history of neurological disease of central origin including seizures; 4) cardiovascular disease including high blood pressure and/or other medical conditions, including metabolic, endocrinological, oncological or autoimmune diseases, and infectious diseases common in iOUD (including Hepatitis B and C or HIV/AIDS); 5) metal implants or other MR contraindications (e.g., claustrophobia); and 6) women who were pregnant or lactating. To recruit a participant sample most representative of OUD in the real world, iOUD were not excluded for a DSM-5 diagnosis of a substance use disorder other than opiates, as long as opiates were the primary drug of choice and/or reason for treatment. Healthy controls (HC) were excluded if they met DSM-5 criteria for a substance use disorder; testing positive for drugs was also exclusionary. A comprehensive diagnostic interview, encompassing the Mini International

Neuropsychiatric Interview[2] (7th ed.) and the Addiction Severity Index[3] (5th ed.), was performed to assess DSM-5 criteria for major psychiatric and substance use disorders.

At baseline, depression and anxiety severity were measured using Beck's Depression and Anxiety Inventories[4,5], respectively; nicotine dependence was measured with the Fagerström Test for Nicotine Dependence (FTND)[6]; alcohol dependence was measured with the Short Michigan Alcoholism Screening Test (SMAST)[7]; and heroin dependence, withdrawal, and craving were evaluated via the Severity of Dependence Scale (SDS)[8], the Subjective Opiate Withdrawal Scale (SOWS)[9], and the Heroin Craving Questionnaire (HCQ; modified from Cocaine Craving Questionnaire[10]), respectively. These measures are summarized for the neurofeedback and peer reference sample in Tables 1 and S1, respectively.

### Offline Neurofeedback Scores

To verify that the neurofeedback learning effect observed in the real-time data also occurs in the fully preprocessed data, we replicated the real-time scoring pipeline offline on each participant's preprocessed 4D BOLD time series (see Methods sections *Functional preprocessing for offline analyses* and *Custom preprocessing and parcellation for offline analyses*). Each run was processed volume-by-volume to simulate the temporal order of the real-time pipeline. A voxel-wise running z-score was computed using Welford's algorithm[11], applied exclusively to movie-content volumes. Volumes preceding the movie onset (preprocessed index 12, equivalent to acquisition TR 23 after accounting for the 20-TR onset delay, 2-TR hemodynamic response lag, and 10 volumes removed during preprocessing) and volumes falling within inter-scene feedback pauses were excluded from the running statistics, matching the real-time pipeline.

At each volume, the z-scored data were multiplied by the pre-trained neurofeedback mask and summed to produce a scalar transformed signal value. These values were accumulated within the predefined station windows, with temporal boundaries adjusted for each scene's position to account for cumulative feedback-pause offsets. At the end of each scene, the accumulated signal was scored using the same correlation-based classifier applied during real-time feedback. Within subjects, real-time and offline scores across stations within runs were significantly correlated ($t(19) = 8.46$, $p < .001$, $r = 0.36$), demonstrating convergence between both sets of scores.

We then applied the same linear mixed-effects model that was applied to the real-time scores (see Methods section *Neurofeedback performance*) to test for a learning effect in the offline scores. For runs 1-4, the real group showed a significant linear increase in neurofeedback performance across runs (Figure S3; $\beta = 0.30$, $t(56.9) = 2.13$, $p = .019$, $d = 0.57$). The sham group showed no such increase ($\beta = -0.19$, $t(56.9) = -1.37$, $p = .912$, $d = -0.36$). The group × run interaction was significant and in the predicted direction (real > sham; $\beta = -0.25$, $t(56.9) = -2.47$, $p = .008$, $d = -0.66$), and groups did not differ at baseline ($\beta = 0.18$, $t(24.5) = 0.94$, two-sided $p = .355$, $d = 0.38$). The larger degrees of freedom in this model reflect a singular fit; the by-subject random slope for run was estimated at zero in the offline data, so the run effect is evaluated against residual variance rather than between-subject slope variability. The corresponding interaction in the real-time scores was weaker and fell short of significance ($\beta = -0.15$, $t(18) = -1.30$, $p = .105$, $d = -0.61$; see *Neurofeedback learning and associated improvements in craving and affect*). Recomputing scores offline on the preprocessed data therefore recovers a group difference in learning during the run 1-4 window that the minimal processing available in real time may have obscured. When all eight runs were included, the linear increase in the real group was no longer significant ($\beta = 0.17$, t(18.0) = 1.33, p = .101, d = 0.63), and the group × run interaction was likewise non-significant ($\beta = -0.04$, t(18.3) = −0.39, p = .352, d = −0.18).

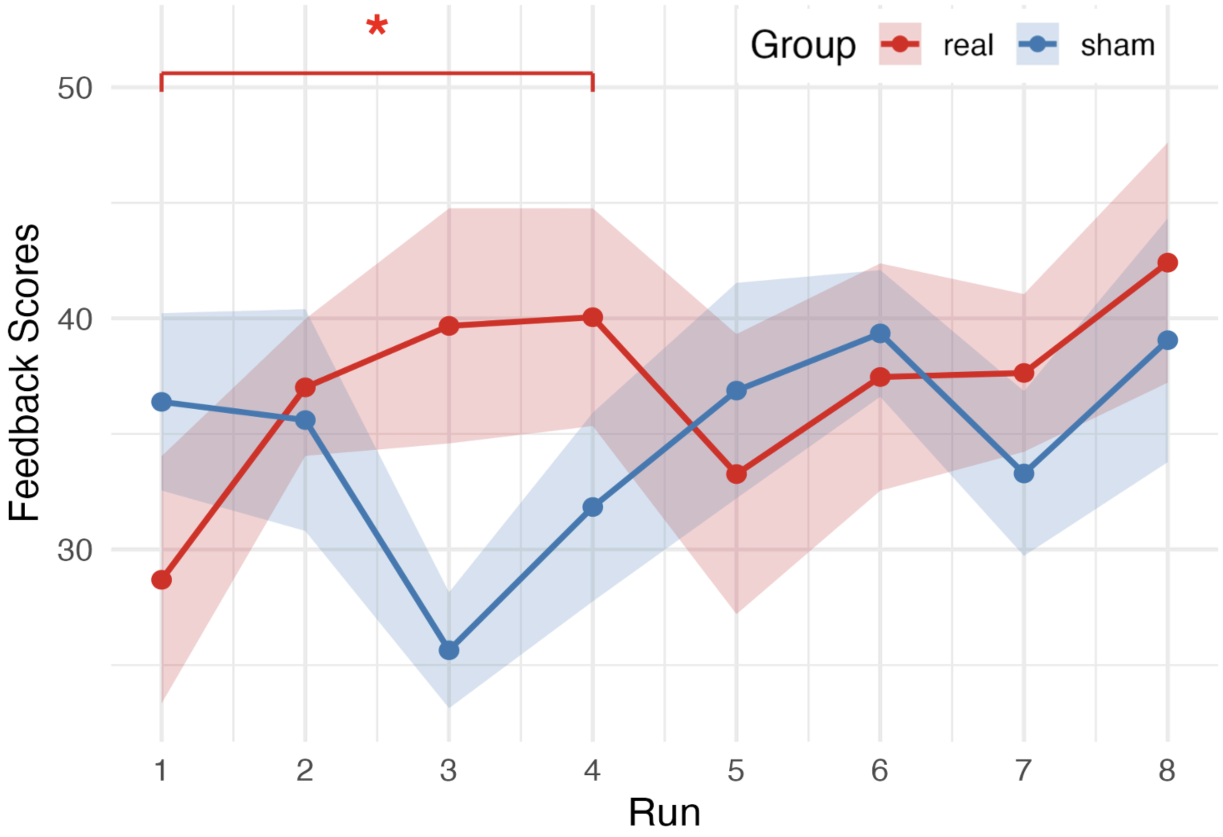


**Figure S3. Post-hoc neurofeedback scores across runs by group.** Mean feedback scores (± standard error) for the real (red, n = 10) and sham (blue, n = 10) neurofeedback groups across eight runs of the task, computed post-hoc using the preprocessed BOLD data. Brackets indicate a significant increase in neurofeedback performance across runs 1-4 in the real group; the non-significant sham group effect is not visualized. The significant group x run effect is also not visualized. *$p < .05$.

### Neurofeedback Strategies

To characterize the cognitive strategies participants employed during PG-MovieNF training, free speech interviews were audio-recorded after each session and transcribed using Pyannote speaker-diarization-3.1 for speaker segmentation and OpenAI Whisper (medium.en) for speech-to-text with word-level timestamps. Words were assigned to speakers by matching temporal midpoints to diarization segments, yielding time-stamped, speaker-labeled transcripts. Strategies were classified into 13 categories defined a priori (e.g., self-talk/affirmation, imagining negative consequences of use, thinking of family/loved ones; see Table S3 for full definitions). Each transcript was coded independently by both a human rater and an LLM (Claude, Anthropic; model claude-sonnet-4-20250514), with identical instructions and category definitions provided to the model, which returned a binary assignment (present/absent) for each category. Disagreements were reviewed and resolved by the human rater to produce the final categorizations.

| Category | Definition |
|---|---|
| Self-talk and affirmation | Using internal or verbal self-directed statements to motivate, reassure, or encourage themselves. Includes repeating mantras, telling themselves they can succeed, expressing pride in progress, or affirming they are glad to no longer be using. |
| Negative consequences | Reflecting on the harmful outcomes of drug use, such as health problems, legal trouble, homelessness, overdose, or the general negativity surrounding substance use. Focuses on why drug use is undesirable. |
| Family and loved ones | Thinking about family members or loved ones (e.g., children, spouse, parents) as a source of motivation. May involve wanting to be present for family or thinking about how drug use affects relationships. |
| Imagine self in scene | Mentally placing themselves into the non-drug scenes depicted in the movie (e.g., playing soccer, being at the park). Involves active visualization of participating in positive activities shown on screen. |
| Recovery and sobriety progress | Drawing on their history of sobriety or recovery, such as reflecting on years of clean time, current program participation, or the fact that they are no longer using substances. |
| Reframe as fiction | Reminding themselves that the movie content is not real (e.g., that the actors are performing, the drugs are props, and the scenes are staged). Cognitive reappraisal that reduces emotional engagement with drug cues. |
| Distraction | Deliberately redirecting attention away from the drug-related content by focusing on background details, thinking about unrelated topics, mentally listing items (e.g., planets), zoning out, or blocking the movie from their awareness. |
| Future goals | Envisioning their future sober self (e.g., imagining getting a job, becoming successful, living a healthier lifestyle, or what life will look like if they stay on the path of recovery). |
| Relate personal experience | Connecting the movie content to their own life history — recalling times they were in similar situations, comparing their past to what is shown on screen, or noting personal relevance of the scenes. |
| Humor and minimizing | Using humor, mockery, or belittlement to reduce the impact of drug scenes (e.g., calling actors bad, making jokes about scenes, thinking of comedy, etc.) |
| Prayer, meditation, and grounding | Employing spiritual or mindfulness-based techniques such as prayer, meditation, grounding exercises, deep breathing, or centering to regulate their response to drug cues. |
| Positive memories | Recalling specific happy or positive memories from their life (e.g., good times with friends, enjoyable activities, pleasant experiences) as a way to counteract negative emotional responses to drug scenes. |
| Reframe negative to positive | Actively taking a negative or drug-related scene and mentally transforming it into something positive or constructive. E.g., imagining how they would change the situation, finding a positive angle, or replacing the negative imagery with a positive alternative. |

**Table S3. Neurofeedback strategy definitions.** The 13 cognitive strategy categories defined a priori for coding post-session free-speech interviews.

For each participant, the proportion of sessions in which each strategy was endorsed was computed. Given the limited sample size, analyses focused on the five most prevalent strategies across participants (family/loved ones, reframe as fiction, recovery/sobriety progress, relate personal experience, and distraction), entered as dependent variables in a one-way MANOVA with group (real vs. sham) as the between-subjects factor. A significant omnibus effect was followed up with univariate ANOVAs for each strategy.

The real and sham groups differed in their overall strategy profile (MANOVA, $F(5,14) = 3.05$, $p = .045$). Univariate follow-ups revealed this was driven by distraction, which the sham group endorsed in a higher proportion of sessions than the real group (sham: $M = 0.41$, real: $M = 0.02$; $F(1,18) = 12.24$, $p = .003$). No other strategy differed significantly between groups (all $F(1,18) \leq 1.51$, all $p \geq .24$; see Figure S4A).

To test whether strategy use changed over training, logistic regression models were fit for each of the five most prevalent strategies with run, group, and their interaction as predictors; p-values were Bonferroni-corrected. Analyses were conducted for runs 1 and 4 ($n = 16$, excluding four participants for whom these runs fell within the same session) and runs 1 and 8 ($N = 20$). No strategy showed a significant run × group interaction (runs 1 and 4: all $|z| \leq 1.29$, all $p \geq .78$; runs 1 and 8: all $|z| \leq 0.93$, all $p = 1.00$).

To characterize the co-occurrence structure among all 13 strategies, pairwise partial Pearson correlations between strategy proportion scores were computed across all participants ($N = 20$), controlling for number of sessions completed, using the *ppcor* package in R. Agglomerative hierarchical clustering (complete linkage) was performed on the dissimilarity matrix ($1 - r$).

Clustering revealed three strategy families (see Figure S4B). The first, *Disengagement*, comprised distraction, humor/minimizing, and positive memories—strategies characterized by redirecting attention away from the movie through avoidance, minimization, or substitution of unrelated positive imagery. The second, *Self-referential reappraisal*, comprised family/loved ones, relate personal experience, and reframe negative to positive—strategies involving relating the movie to participants' personal lives and reframing specific moments. The third, *Goal-directed self-regulation*, comprised future goals, prayer/meditation/grounding, self-talk/affirmation, recovery/sobriety progress, negative consequences, imagine self in scene, and reframe as fiction—strategies reflecting deliberate deployment of motivational and regulatory resources, including recovery-oriented thinking, somatic grounding, and effortful perspective-taking.

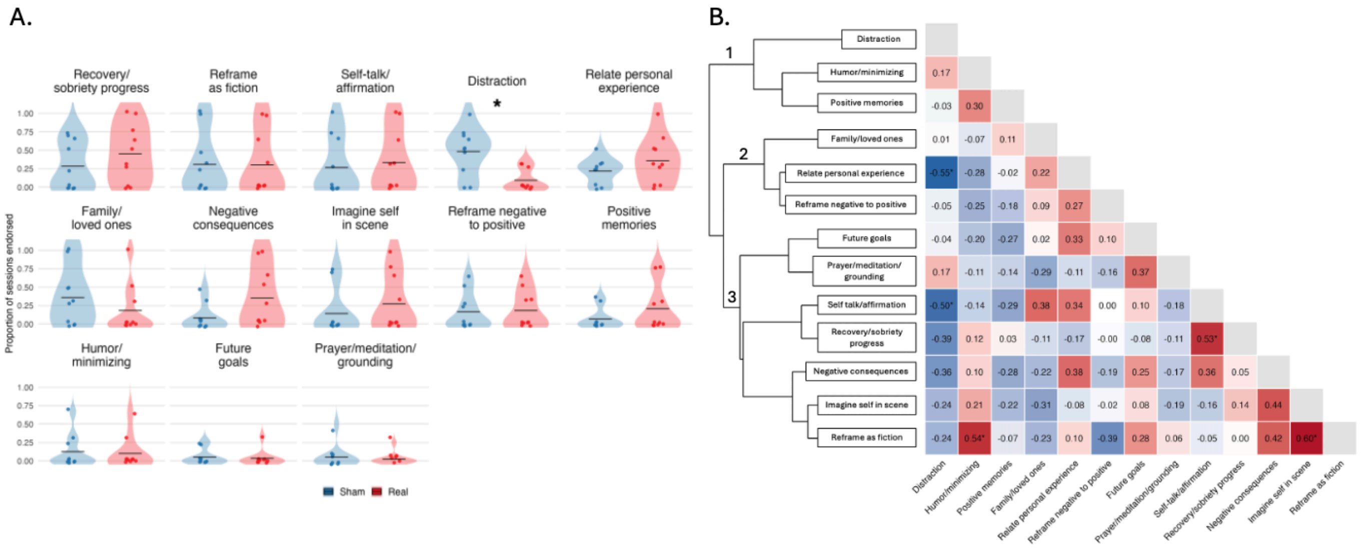


**Figure S4. Cognitive strategy profiles during neurofeedback training.** Cognitive strategy usage profiles throughout PG-MovieNF training. **A.** Strategy endorsement by group. Violin plots show the distribution of the proportion of sessions in which each of the 13 *a priori* strategy categories was endorsed for the sham (blue) and real (red) groups; points denote individual participants and horizontal lines denote group means. Group differences were tested for the five most prevalent strategies (MANOVA followed by univariate ANOVAs); the sham group endorsed distraction in a higher proportion of sessions than the real group. **B.** Co-occurrence structure among strategies. Lower-triangular matrix of pairwise partial Pearson correlations between strategy endorsement proportions across all participants, controlling for the number of sessions completed. Warm colors indicate positive and cool colors indicate negative correlations; cell values denote the partial r, with asterisks marking correlations significant at $p < .05$. The dendrogram (left) shows agglomerative hierarchical clustering (complete linkage) on the dissimilarity matrix ($1 - r$), which identified three strategy families: (1) Disengagement (distraction, humor/minimizing, positive memories), (2) Self-referential reappraisal (family/loved ones, relate personal experience, reframe negative to positive), and (3) Goal-directed self-regulation (future goals, prayer/meditation/grounding, self-talk/affirmation, recovery/sobriety progress, negative consequences, imagine self in scene, reframe as fiction). $^*p < .05$.

As an exploratory analysis, we examined whether endorsement frequency of the three most prevalent strategies across participants (relate personal experience, recovery/sobriety progress, and distraction) was associated with neurofeedback performance. An OLS regression was fit with mean neurofeedback score as the outcome and these endorsement proportions as standardized predictors.

The model approached significance (adjusted $R^2 = .237$, $F(3,16) = 2.97$, $p = .063$). Among individual predictors, relating scenes to personal experience was negatively associated with neurofeedback performance ($\beta = -0.61$, $p = .024$), whereas recovery/sobriety progress ($\beta = +0.12$, $p = .620$) and distraction ($\beta = -0.21$, $p = .423$) were not significantly associated.
